# Prognostic models predicting transition to psychotic disorder using blood-based biomarkers: a systematic review and critical appraisal

**DOI:** 10.1101/2023.09.08.23295245

**Authors:** Jonah F. Byrne, David Mongan, Jennifer Murphy, Colm Healy, Melanie Fӧcking, Mary Cannon, David R. Cotter

## Abstract

**Background:** Accumulating evidence suggests individuals with psychotic disorder show abnormalities in metabolic and inflammatory processes. Recently, several studies have employed blood-based predictors in models predicting transition to psychotic disorder in risk-enriched populations. A systematic review of the performance and methodology of prognostic models using blood-based biomarkers in the prediction of psychotic disorder from risk-enriched populations is warranted.

**Methods:** Databases (PubMed, EMBASE and PsycINFO) were searched for eligible texts from 1998 to 15/05/2023 which detailed model development or validation studies. The checklist for Critical Appraisal and Data Extraction for Systematic Reviews of Prediction Modelling Studies (CHARMS) was used to guide data extraction from eligible texts and the Prediction Model Risk of Bias Assessment Tool (PROBAST) was used to assess risk of bias and applicability of the studies. A narrative synthesis of included studies was performed.

**Results:** 17 eligible studies were identified: 16 eligible model development studies and one eligible model validation study. A wide range of biomarkers were assessed including nucleic acids, proteins, metabolites and lipids. The range of C-index (area under the curve) estimates reported for the models was 0.67-1.00. No studies assessed model calibration. According to PROBAST criteria, all studies were at high risk of bias in the analysis domain.

**Discussion:** While a wide range of potentially predictive biomarkers were identified in the included studies, most studies did not account for overfitting in model performance estimates, no studies assessed calibration, and all models were at high risk of bias according to PROBAST criteria. External validation of the models is needed to provide more accurate estimates of their performance. Future studies which follow the latest available methodological and reporting guidelines and adopt strategies to accommodate required sample sizes for model development or validation will clarify the value of including blood-based biomarkers in models predicting psychosis.

## Introduction

### Background and Rationale

Recent research in the field of early intervention in psychosis has focused on building models to predict the development of psychotic disorder [1–3]. These models have largely been developed in “clinical high-risk” populations, which include individuals showing prodromal symptoms or genetic risk combined with functional decline, as determined with validated assessment tools such as the Comprehensive Assessment of At-Risk Mental State (CAARMS) [4] and the Structured Interview for Prodromal Symptoms (SIPS) [5,6]. Meta-analytic estimates indicate 19% of individuals at clinical high-risk develop psychosis within two years [7].

Accumulating evidence points towards abnormalities in metabolic and inflammatory processes in individuals with psychosis [8–10] and there is some evidence to suggest that these abnormalities may precede medication use [9] or even the onset of psychosis [11,12]. The prognostic value of peripheral markers over lifestyle or environmental factors, such as smoking and exercise, is unclear [13,14]. However, several studies have employed a range of blood-based predictors in models predicting transition to psychotic disorder, with several published since the last systematic reviews of models in the field [15–19]. A recent large-scale systematic review included all prediction models in psychiatry, except those using biological predictors [20]. Biological predictors have the advantage of often being more objective and precise than, for example, scores given on symptom scales. Blood biomarkers are among the least invasive biological parameters, with relative low-cost. There is a clear pathway for their integration into clinical practice, as they could be measured along with routine blood markers, or predictors could consist of routine blood measures. As such, a systematic review of prognostic models predicting transition to psychotic disorder using blood-based biomarkers is warranted. Furthermore, the weak standard of prediction modelling methodology in psychiatry and medicine in general has been highlighted previously, in particular regarding the need for validation and implementation of models [20–22]. Therefore, a review of methodology that takes into account the interaction between the use of blood-based biomarkers and prediction modelling may help future research in the field.

### Objectives

To systematically review the performance and methodology of models predicting transition to psychotic disorder from risk-enriched populations, with a focus on those using blood-based biomarkers, to determine their potential utility in predictive models and to help guide future research in the field.

## Methods

This review was reported using the guidelines for transparent reporting of multivariable prediction models for individual prognosis or diagnosis: systematic reviews and meta-analyses (TRIPOD-SRMA) [23].

### Eligibility Criteria

Studies were eligible for inclusion if they; 1) described the development, validation or updating of a prognostic model of transition to psychotic disorder in “at-risk” (or similar psychosis risk-enriched populations) help-seeking individuals, 2) used blood-based biomarkers in the prognostic model described, 3) were published in peer-reviewed journals, 4) were published after 1998 (after the first prospective studies using clinical high-risk criteria were published; [24]) and 5) had full-texts available in English. The PICOTS (Population, Intervention, Comparator, Outcome, Timing, Setting) criteria were used to guide the development of eligibility criteria and can be found in the Supplementary Material.

Studies were excluded if they focused on psychiatric disorders other than psychotic disorders (for example studies solely focusing on prediction of depression) and if they investigated risk factors or longitudinal associations but did not report the development and/or validation of a risk prediction model. Studies that reported only on models including blood-based predictors in combination with brain or cerebrospinal fluid biomarkers were excluded as the predictive value of the blood-based biomarkers in the models would be difficult to determine precisely, and as the aim of the review was to identify models that required the measurement of blood biomarkers only, without the requirement for a further invasive and expensive procedure. Studies must have had a binary outcome of transition to psychosis to be included in the current study (studies predicting continuous outcomes were excluded, for example psychotic symptom scales). Studies of continuous outcomes were excluded for several reasons, including a) transition is the key outcome in the literature, b) the difficulty in implementing a prediction model in clinical practice without a clear diagnostic outcome, c) individual clinical scales (positive symptoms or negative symptom scales) can’t define diagnosis alone, d) potentially multiple different outcomes (functioning and clinical symptom subscales) that may use multiple different non-comparable scales (e.g. the Positive And Negative Symptom Scale and the Community Assessment of Psychic Experiences).

### Information Sources and Search Strategy

PubMed, EMBASE and PsycINFO were searched from 01/01/1998 to 15/05/2023 using the following general search strategy: psychosis risk-enrichment keywords AND transition to psychotic disorder keywords AND prediction modelling keywords AND blood-based biomarker keywords. The search strategy was developed with the use of established strategies for searching for predictive modelling studies [25]. The full search strategies, as formatted for each database, are included in the Supplementary Material.

In an attempt to find other references that may meet inclusion criteria, reference lists of relevant reviews that appeared in the databases searched were examined and forward citation searching was carried out on Google Scholar for texts eligible for full-text screening up to 01/06/2023. Where clearly eligible models were detailed in conference abstracts and corresponding full-texts could not be found, we contacted the corresponding authors for information on potential unpublished full-texts.

### Selection Process

Duplicate records were identified with guidance from previous recommendations [26] and removed. Abstracts identified by the search strategy were screened independently by JFB, DM and JM. Prediction modelling studies that clearly did not meet the eligibility criteria were excluded and the full-texts of all other studies were examined. Disagreements were resolved through discussion and or by referral to a third author (DRC).

### Data Collection Process

Data were extracted from studies using the CHARMS checklist for Critical Appraisal and Data Extraction for Systematic Reviews of Prediction Modelling Studies [27]. Data were extracted independently by JFB, DM and JM. Disagreements were resolved through discussion or referral to a third author (CH or MF). Where several similar models were presented in studies (e.g. models with minor differences in predictors included), data pertaining to the final model as indicated by the study authors was extracted. Where study authors did not indicate a final model, data pertaining to the best performing model was extracted.

As part of the CHARMS, we extracted two main model performance metrics: discrimination (how well the model differentiates between individuals who do and do not develop the outcome; the Concordance (C)-index [28], which is equivalent to the area-under-the-curve in the case of a binary outcome) and calibration (how well the predicted probabilities match the actual proportion of outcomes) [29], where available. Where studies reported on the added predictive value of blood-based biomarkers to models, relevant metrics and tests (such as the likelihood ratio chi-square test) of added predictive information were extracted, if available.

### Risk of Bias and Applicability Assessment

Risk of bias assessments were carried out using the prediction model risk of bias assessment tool (PROBAST) [30,31], which assesses risk of bias in the selection of participants, measurement of predictors or outcomes and in the analysis. Using the PROBAST tool, concerns of applicability of the study to the review question were also rated in the domains selection of participants, measurement of predictors or measurement of outcomes. PROBAST ratings for risk of bias and applicability assessments can be either high, low or unclear. Assessments were carried out independently by JFB, DM and JM. Disagreements were resolved through discussion with a third author (CH). PROBAST figures were generated using the robvis package [32] in R (https://github.com/mcguinlu/robvis). Required sample sizes for precise estimates of model performance on external validation were calculated in R (https://github.com/c-qu/samplesize-validation) according to guidelines from a previous publication [33].

### Synthesis Methods

We planned to use a narrative synthesis method [34]. We stratified performance estimates of the included model development studies based on the PROBAST signalling question “Were model overfitting and optimism in model performance accounted for?”. As per PROBAST guidance, model overfitting and optimism in model performance estimates were accounted for if both internal validation and shrinkage techniques were applied, and if predictor selection procedures (e.g. univariable screening or backwards selection) were included in the internal validation framework [31]. Meta-analyses were to be carried out only if a particular model had multiple external validation studies [35].

## Results

### Study Selection

A PRISMA flow diagram is presented in Figure 1. The database search identified 2,565 records. Following removal of duplicates, 1,677 titles and abstracts were screened, and 45 articles were brought forward for full-text screening. 13 records (relating to 13 studies) were included, and 32 records were excluded. Reasons for exclusion can be summarised in the following categories: not predictive modelling, no blood-based predictors used, ineligible study design (participants or outcome do meet eligibility criteria), conference abstract only. We contacted authors of three conference abstracts which detailed clearly eligible models for further information on potential full-texts related to the abstracts. Authors of two conference abstracts confirmed that both abstracts related to a single study for which we had already obtained a peer-reviewed full-text. We did not receive a response from the authors of one conference abstract which detailed a clearly eligible model.

**Figure 1:**
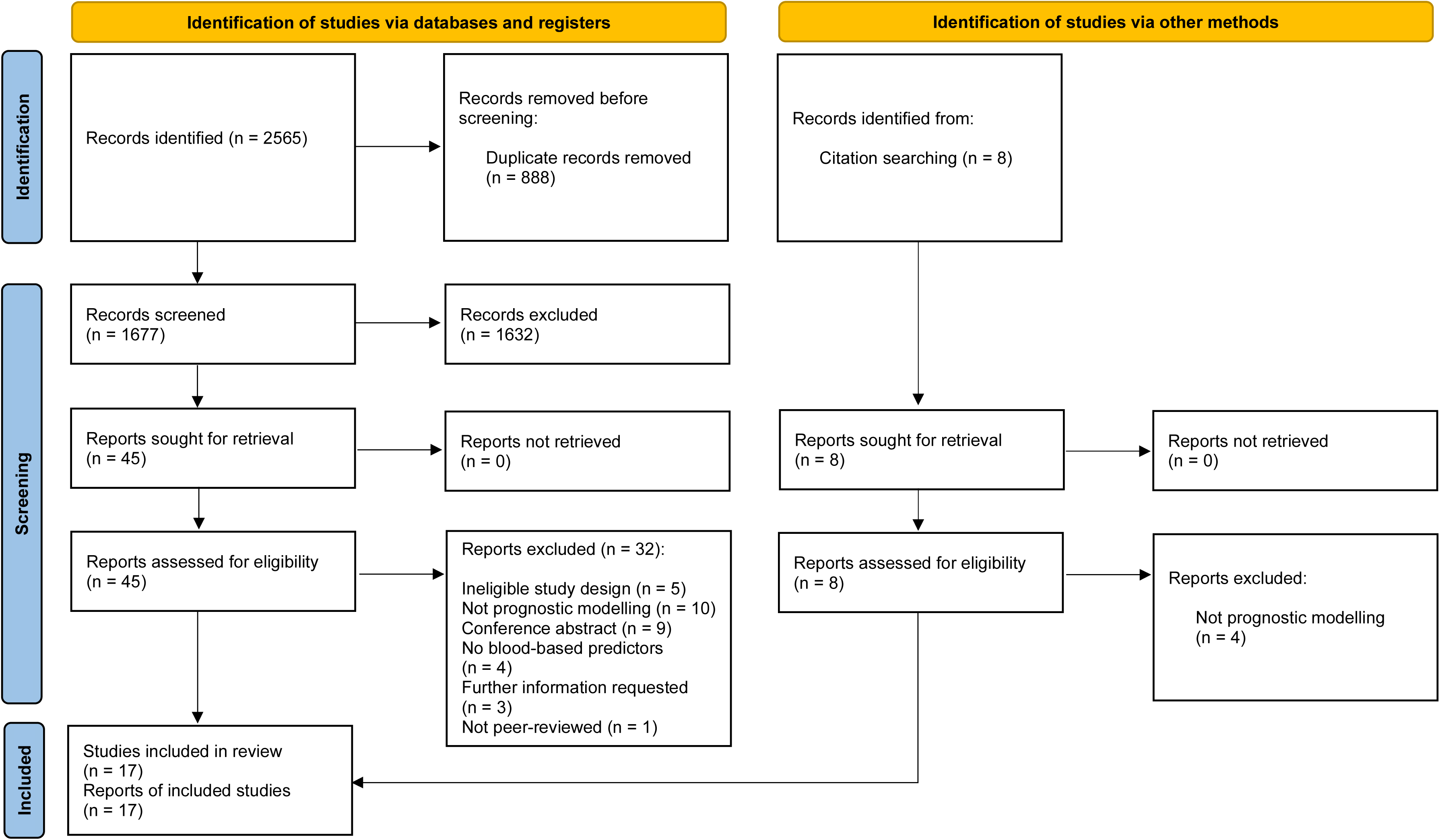
PRISMA flow diagram of study selection.

Forward citation searching of the 45 full-texts identified eight further potentially eligible full-texts, of which four were excluded (not predictive modelling) and four records (relating to four studies) were included (two of which were full-text reports of conference abstracts identified in the database search). Therefore, 17 studies were included overall.

### Study Characteristics

Of the 17 included studies, 16 were prognostic model development studies [15–19,36–46] and one study was a prognostic model external validation study [47]. All studies were conducted in outpatients. There were five models developed in the Shanghai At Risk for Psychosis (SHARP) study [18,19] and the extension of that study [44–46]. There were three model development studies each for the North American Prodrome Longitudinal Study (NAPLS 2) cohort [36,38,41] and for the EU Gene-Environment Interaction (EU-GEI) study [15,16,42]. Two studies were developed models in participants of the Vienna omega-3 randomised-controlled trial [37,39]. There was one model developed in the Personalised Prognostic Tools for Early Psychosis Management (PRONIA) study [17], one model developed in a Korean cohort study [40], one model developed in participants recruited from the Outreach and Support in South London high-risk service [43], and one validated in the ICAAR (Influence of Cannabis in the emergence of psychopathological symptoms in Adolescents and Adults at-Risk) study [47].

Mean study participant ages ranged from 15.8 years to 24.6 years. The majority of studies (16/17) defined participants at increased risk of psychosis through use of the SIPS (ten studies), CAARMS (four studies) or CAARMS-equivalent (two studies) criteria. A wide range of biomarkers were assessed including cytokines (four studies), single-nucleotide polymorphisms (SNPs; four studies), hormonal, inflammatory and metabolic-related analytes (two studies), ribonucleic acids (two studies), lipids (two studies), proteins (one study), metabolites (one study), and glutathione (one study) (Table 1).

**Table 1:**
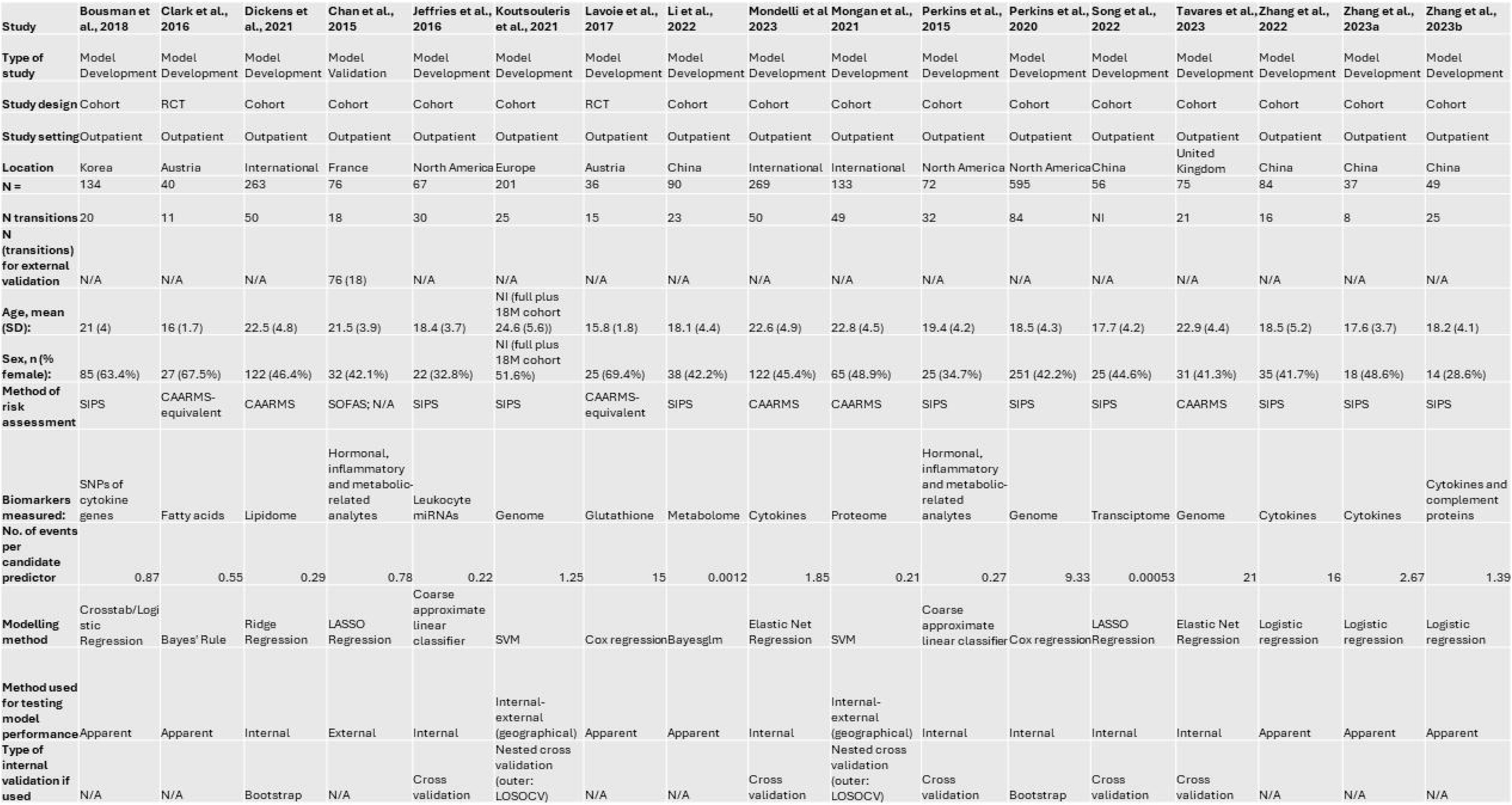
Study Characteristics. Structured Interview for Psychosis Risk Syndromes (SIPS), Comprehensive Assessment of At-Risk Mental State (CAARMS), Social and Occupational Functioning Assessment Scale (SOFAS), micro-ribonucleic acid (miRNA), single-nucleotide polymorphisms (SNPs), least-absolute shrinkage and selection operator (LASSO), support vector machine (SVM), bayesian generalised linear model (bayesglm), logistic regression (LR), leave one site out cross-validation (LOSOCV).

Of the 16 development studies, nine internally validated model performance, of which five accounted for optimism in their performance estimates [15,17,41]. Four studies which used internal validation did not include the predictor selection process within the internal validation procedure. The remaining seven studies reported apparent performance (Table 1). Reported C-indices for logistic regression models ranged from 0.67-1.00. Reported C-indices for cox models ranged from 0.82-0.88. One study reported a balanced accuracy of 46.2%. None of the studies reported calibration measures or assessed clinical utility of their models. Further characteristics and performance metrics of the included studies are detailed in the Supplementary Material.

### Risk of Bias and Applicability Assessments of Included Studies

All studies included in the systematic review were at high overall risk of bias according to PROBAST criteria. This was related to risk of bias in the analysis domain (Figure 2; Supplementary Table 3).

**Figure 2:**
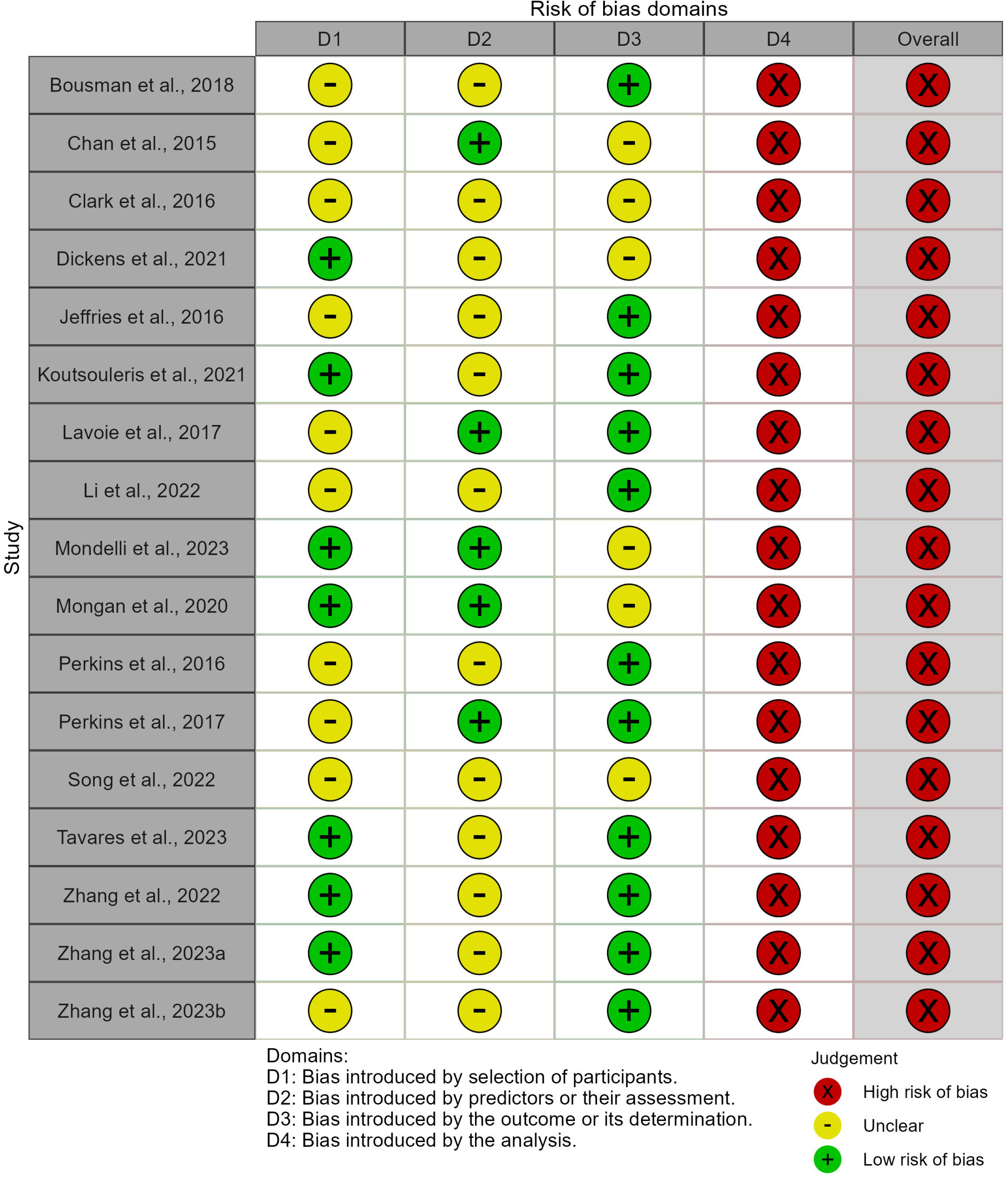
Results of risk of bias assessments in each domain.

The risk of bias due to the selection of participants was rated unclear in ten out of 17 studies (58.8%). Generally, this related to either a lack of reporting of exclusion criteria used, a lack of comparison between included and excluded participants, or a lack of reporting of how many exclusions were made due to potentially inappropriate criteria (such as abnormal levels of blood parameters). Regarding applicability concerns in this domain, for the same reasons it was unclear in ten studies (58.8%) whether participants matched the review question (Figure 3).

**Figure 3:**
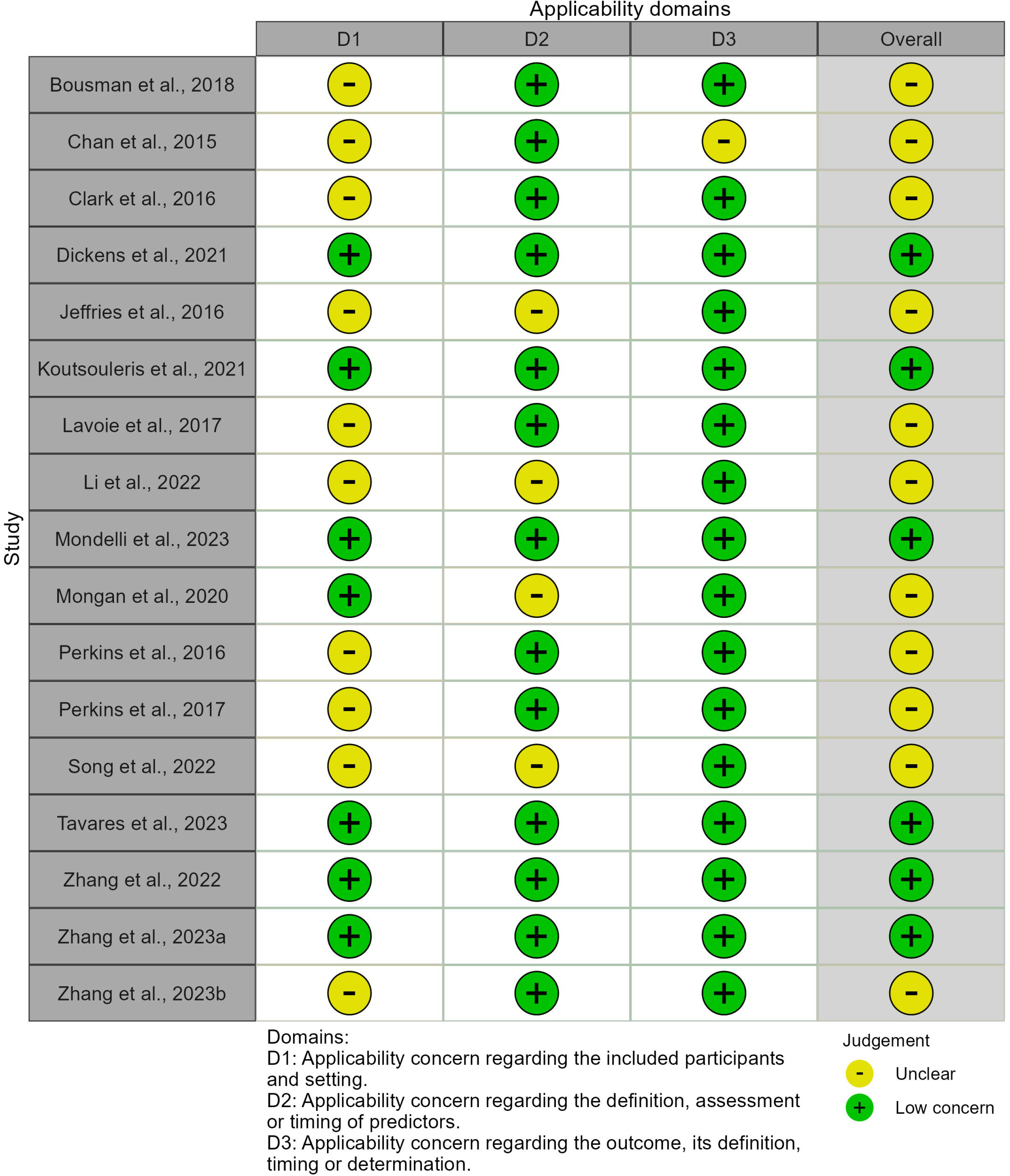
Summary of applicability concerns in each domain.

Risk of bias due to the predictors or their assessment was unclear for 12 out of 17 studies (70.6%) as they did not report whether predictor assessments were made blind to the outcome data. The method of predictor assessment used in four studies (23.5%) may not match the review question, as it was unclear if the biomarker measurement methods used provide absolute quantification. A lack of standardised units for the biomarker measurement could hinder the generalisability of models. There were low applicability concerns for the remaining 13 studies in this applicability domain.

Risk of bias introduced by the outcome or its determination was low for 11 studies which used standard measures or structured interviews. Risk of bias introduced by the outcome was unclear for six studies (35.3%), largely because insufficient information on how the outcome was determined was reported. There were generally low concerns regarding the applicability of the outcome, with one study rated unclear in this domain, as the timing of the outcome determination relative to collection of the blood sample was unclear.

All studies were at high risk of bias in the analysis domain. None of the studies reported measures of model calibration. Generally, studies did not have a sufficient number of participants with the outcome or had a low number of events per candidate predictor. 9/16 development studies (56.3%) had less than one event per candidate predictor, and 14/16 (87.5%) had less than three events per candidate predictor. The one model validation study had a sample size of 76. However, we calculated that with the given study outcome prevalence of 0.237 and expected external validation C-index of 0.80, the minimum recommended sample size for precise estimation of model performance metrics on external validation would be 709. [33,48,49]. 5/16 (31.3%) development studies accounted for model overfitting and optimism in performance estimates. Four of these studies used cross-validation for internal validation and penalised regression models to shrink coefficients. One of these studies used the bootstrap for internal validation and shrinkage. Other limitations included not accounting for subsampling of controls, not reporting how missing data was handled or the use of univariable analysis to select predictors for inclusion in the model.

### Narrative Synthesis

None of the included studies assessed model calibration, a key metric for which a model must perform acceptably if it is to be used to determine whether an intervention should be offered [29]. The logistic regression model C-indices in model development studies which did not account for optimism in their performance estimates ranged from 0.81 to 1 (with one study reporting a cox model C-index of 0.82). Model development studies which accounted for optimism in their performance estimates reported C-indices from logistic regression models ranging from 0.67-0.95 (with one study reporting a cox model C-index of 0.88 and one study reporting a balanced accuracy of 46.2%).

Three studies which adjusted their performance estimates for optimism used polygenic risk scores (PRS) as a blood-based predictors [17,41,43]. Perkins et al. [41] investigated adding PRS to an established cox regression model comprised of clinical predictors. When PRS was included as a variable together with the clinical predictors, there was no evidence from a likelihood ratio chi-square test that the PRS added predictive value. Furthermore, the C-index was unchanged for participants of non-European descent and was similar for participants of European descent (point estimate increase of 0.01). In line with this, Koutsouleris et al. [17] found that a model based on PRS variables had a similar C-index to that achieved by predictions from clinical raters, and Tavares et al. [43] found that a PRS model did not discriminate between individuals who transitioned and did not transition better than chance.

The final model presented by Chan et al. [47] and the “15-analyte” model in Perkins et al. [36] had two overlapping predictors, Interleukin-8 and Factor VII. While model coefficients were not available in Perkins et al. [36], the two studies presented the same direction of effect for IL-8 and opposite directions of effect for Factor VII in univariate analyses. Zhang et al. [46] and Mondelli et al [42] did not find evidence for the predictive ability of IL-8 for transition to psychotic disorder. Zhang et al. 2022 [44], Zhang et al. 2023a [45] and Mondelli et al. [42] all included different cytokine ratios in their models (IL-1β/IL-6, IL-2/IL-6 and IL-10/IL-6 respectively) with the aim of capturing inflammatory balance. Chan et al. [47] and Mongan et al. [15] both included Alpha-2-Macroglobulin as a predictor in their final models, though coefficients were in opposite directions. No other overlapping predictors were noted between final models presented in studies.

## Discussion

We undertook a focused systematic review of models predicting transition to psychosis with use of recent guidelines for systematically reviewing prognostic models [31,50]. Models developed with blood biomarkers require unique consideration, as any predictive benefit of blood biomarkers over clinical predictors must outweigh the disadvantages associated with their measurement, namely the cost or potential lack of wide accessibility. While studies included in this review described a wide range of blood biomarkers that potentially have altered concentrations preceding psychosis, the prognostic models including blood biomarkers were not developed according to the latest methodological recommendations, lacked calibration and lacked sufficient external validation. Therefore, similar to recent systematic reviews of prediction models in psychiatry [20,51,52], we did not find evidence for a model suitable for implementation into clinical practice [53].

As all the studies were rated at high risk of bias, we are unable to recommend a particular model to be externally validated, and it is unclear at the present time whether any specific blood biomarkers could potentially contribute to improved prediction of transition to psychosis in individuals at risk. However, evidence from three studies suggested that models including polygenic risk scores do not sufficiently outperform models based on clinical variables or clinical rater predictions. A wide range of other biomarkers were assessed in the included studies, however, going forward, the field will need to externally validate models to truly estimate their generalizability. Risk prediction models cannot be recommended for clinical practice without sufficient external validation.

In general, the reporting of study design and methods could be improved in the field. We recommend that future studies follow the transparent reporting of a multivariable prediction model for individual prognosis or diagnosis (TRIPOD) reporting guidelines [54] and explicitly report on participant exclusion criteria and whether predictors were assessed blind to outcome status. Blood-biomarkers in cohort studies are often assessed after the outcome is determined and therefore studies involving blood-biomarkers are at particular risk of bias in this domain. As well as blinding, studies should report whether samples were randomised prior to biomarker measurement. Recent guidance proposes block randomisation as a gold standard [55,56].

Sixteen out of the 17 studies in this review defined their participants to be at risk of psychosis through use of SIPS and CAARMS interviews for prodromal symptoms or reduced functioning combined with genetic risk. While these established “clinical-high risk” and “at-risk mental state” constructs are risk-enriched populations with an estimated 2-year transition rate of 19% [7], they may only capture a minority (4-14%) of the population who present with a first episode of psychosis, indicating a relatively poor predictive capacity [57–59]. There have been recent calls to expand the clinical high risk paradigm such as to include individuals attending child and adolescent mental health services or presenting to emergency departments due to self-harm, as they are at similar risk of psychosis [60–62]. This “systems approach” may capture a greater proportion of first episode psychosis cases than prodromal constructs. We recommend that future studies are designed in populations with a higher predictive capacity for psychosis [62], as this may help to mitigate issues with insufficient sample sizes in the field and potential recruitment bias.

Some of the studies in this review excluded participants based on established psychiatric diagnoses or when a participant’s prodromal symptoms were caused by a mood or anxiety disorder. In light of studies showing that psychotic symptoms are highly prevalent in disorders of depression and anxiety [63] and that psychosis can be predicted in individuals with non-psychotic mental illnesses [64–66], these may be inappropriate exclusion criteria. On the other hand, a more nuanced assessment of clinical and biological risk factors associated with psychosis, such as minimal self [67], circadian rhythms [68] or trait-like EEG signatures [69], could be used to reduce biological heterogeneity. Reducing biological heterogeneity could complement traditional risk-enrichment approaches and allow for the identification of more replicable blood biomarkers of psychosis risk.

Half of the studies included in this systematic review considered over 100 candidate predictors. With the growing popularity of “omics” methods, it must be highlighted that the latest research does not suggest that data-driven methods of predictor selection involving the outcome data can alleviate overfitting in situations of high dimensionality and low sample size. Univariable screening has long been highlighted as problematic [70]. Multivariable selection methods such as LASSO have recently been shown to be unstable in small sample sizes or with small numbers of events [71]. Researchers should be aware that penalisation methods do not solve the issue of a small ratio of events to predictors. Sample size calculators for the development or validation of prediction models with binary or time-to-event outcomes are now easily accessible and should be utilised prior to designing studies to ensure adequate power [49,72,73].

One of the main limitations of the studies in the analysis domain was the lack of calibration of models. In the first instance, clinical prediction models must produce predicted event probabilities for each individual rather than binary event or non-event predictions alone - for a model to be implemented into clinical practice, a probability cut-off relating to maximum clinical benefit is required to determine whether a specific intervention should be offered or not [22,30,74]. Calibration measures how well the predicted probabilities match the observed proportion of outcomes, i.e. the accuracy of risk estimates, and models can have poor calibration even when models show good discrimination. An over- or underestimation of the probability of developing a psychotic disorder is ethically unacceptable, and would lead to inappropriately offered interventions or undertreatment [29]. Future studies in the field should assess model calibration to improve the chances of models being implemented clinically.

The studies included in this review had further limitations in the analysis domain. Some studies did not handle missing data to PROBAST standards. Missing biomarker data is often related to biomarker concentrations being below the limit of detection. In this case, the data should be considered missing not-at-random. PROBAST guidelines recommend multiple imputation as best practice in prediction modelling [31], and solutions combining multiple imputation with left-censored missing data have been proposed [75]. Furthermore, participant subsampling was frequent. When subsampling is necessary, researchers should weight cases and controls by the inverse of their sampling fractions in analyses [28].

This review has several strengths. The review has benefitted from prospective registration and the use of recommended reporting guidelines [23], search strategies [25], data extraction tools [27] and risk of bias assessment tools [30]. However, there are also several limitations to this review. We were unable to perform meta-analyses as we did not identify any models that were externally validated multiple times. Meta-analyses of models with different predictors or validation approaches would not have been readily interpretable. Due to limitations of the modelling strategies in the studies and lack of external validation of models, we were not able to perform a head-to-head comparison of the performance of each of the prediction models as the performance estimates were at high risk of bias. Finally, we acknowledge that the concept of a binary “transition” to psychosis, even in the presence of assessment criteria, can be subjective or can sometimes represent small increases in the severity or frequency of symptoms [76]. This review did not examine the prediction of positive symptoms as continuous outcomes, which may be worth examining in future reviews in the field.

In conclusion, while there have been several studies developing models using blood-based biomarkers for prediction of transition to psychotic disorder, this review found no models that are ready for implementation in clinical practice, and the value of including blood-based biomarkers in models predicting transition to psychosis is unclear due to the high risk of bias of the eligible studies. The field of prediction modelling is rapidly progressing and it should be noted that new methodological recommendations have been made since the majority of the studies in this review were published [71,72]. Future studies should aim to follow the latest available reporting guidelines, assess model calibration, internally and repeatedly externally validate models, and adopt strategies to accommodate minimum required sample sizes in order to maximise potential clinical benefits and outcomes for patients.

### Registration and Protocol

This systematic review was prospectively registered with PROSPERO, CRD42022302047.

## Data Availability

All data produced in the present work are contained in the manuscript and supplemental files.

## References

1. Riecher-Rössler A, Studerus E. Prediction of conversion to psychosis in individuals with an at-risk mental state: a brief update on recent developments. Current Opinion in Psychiatry. 2017;30:209–219.

2. Sanfelici R, Dwyer DB, Antonucci LA, Koutsouleris N. Individualized Diagnostic and Prognostic Models for Patients With Psychosis Risk Syndromes: A Meta-analytic View on the State of the Art. Biological Psychiatry. 2020;88:349–360.

3. Montemagni C, Bellino S, Bracale N, Bozzatello P, Rocca P. Models predicting psychosis in patients with high clinical risk: a systematic review. Frontiers in Psychiatry. 2020;11:223.

4. Yung AR, Yung AR, Pan Yuen H, Mcgorry PD, Phillips LJ, Kelly D, et al. Mapping the onset of psychosis: the comprehensive assessment of at-risk mental states. Australian & New Zealand Journal of Psychiatry. 2005;39:964–971.

5. Miller TJ, McGlashan TH, Rosen JL, Cadenhead K, Ventura J, McFarlane W, et al. Prodromal assessment with the structured interview for prodromal syndromes and the scale of prodromal symptoms: predictive validity, interrater reliability, and training to reliability. Schizophrenia Bulletin. 2003;29:703–715.

6. Fusar-Poli P, Borgwardt S, Bechdolf A, Addington J, Riecher-Rössler A, Schultze-Lutter F, et al. The Psychosis High-Risk State: A Comprehensive State-of-the-Art Review. JAMA Psychiatry. 2013;70:107–120.

7. Salazar de Pablo G, Radua J, Pereira J, Bonoldi I, Arienti V, Besana F, et al. Probability of Transition to Psychosis in Individuals at Clinical High Risk: An Updated Meta-analysis. JAMA Psychiatry. 2021;78:970–978.

8. Fraguas D, Díaz-Caneja CM, Rodríguez-Quiroga A, Arango C. Oxidative stress and inflammation in early onset first episode psychosis: a systematic review and meta-analysis. International Journal of Neuropsychopharmacology. 2017;20:435–444.

9. Upthegrove R, Manzanares-Teson N, Barnes NM. Cytokine function in medication-naive first episode psychosis: a systematic review and meta-analysis. Schizophrenia Research. 2014;155:101–108.

10. Perry BI, McIntosh G, Weich S, Singh S, Rees K. The association between first-episode psychosis and abnormal glycaemic control: systematic review and meta-analysis. The Lancet Psychiatry. 2016;3:1049–1058.

11. Mongan D, Healy C, Jones HJ, Zammit S, Cannon M, Cotter DR. Plasma polyunsaturated fatty acids and mental disorders in adolescence and early adulthood: cross-sectional and longitudinal associations in a general population cohort. Translational Psychiatry. 2021;11:1– 13.

12. Perry BI, Stochl J, Upthegrove R, Zammit S, Wareham N, Langenberg C, et al. Longitudinal Trends in Childhood Insulin Levels and Body Mass Index and Associations With Risks of Psychosis and Depression in Young Adults. JAMA Psychiatry. 2021;78:416–425.

13. Osimo EF, Baxter L, Stochl J, Perry BI, Metcalf SA, Kunutsor SK, et al. Longitudinal association between CRP levels and risk of psychosis: a meta-analysis of population-based cohort studies. Npj Schizophrenia. 2021;7:31.

14. Firth J, Solmi M, Wootton RE, Vancampfort D, Schuch FB, Hoare E, et al. A meta-review of “lifestyle psychiatry”: the role of exercise, smoking, diet and sleep in the prevention and treatment of mental disorders. World Psychiatry. 2020;19:360–380.

15. Mongan D, Föcking M, Healy C, Susai SR, Heurich M, Wynne K, et al. Development of proteomic prediction models for transition to psychotic disorder in the clinical high-risk state and psychotic experiences in adolescence. JAMA Psychiatry. 2021;78:77–90.

16. Dickens AM, Sen P, Kempton MJ, Barrantes-Vidal N, Iyegbe C, Nordentoft M, et al. Dysregulated lipid metabolism precedes onset of psychosis. Biological Psychiatry. 2021;89:288–297.

17. Koutsouleris N, Dwyer DB, Degenhardt F, Maj C, Urquijo-Castro MF, Sanfelici R, et al. Multimodal machine learning workflows for prediction of psychosis in patients with clinical high-risk syndromes and recent-onset depression. JAMA Psychiatry. 2021;78:195–209.

18. Li Z, Zhang T, Xu L, Wei Y, Cui H, Tang Y, et al. Plasma metabolic alterations and potential biomarkers in individuals at clinical high risk for psychosis. Schizophrenia Research. 2022;239:19–28.

19. Song W, Xu L, Zhang T, Wang W, Fu Y, Xu Q, et al. Peripheral transcriptome of clinical high-risk psychosis reflects symptom alteration and helps prognosis prediction. Psychiatry and Clinical Neurosciences. 2022. 2022.

20. Meehan AJ, Lewis SJ, Fazel S, Fusar-Poli P, Steyerberg EW, Stahl D, et al. Clinical prediction models in psychiatry: a systematic review of two decades of progress and challenges. Molecular Psychiatry. 2022;27:2700–2708.

21. Studerus E, Ramyead A, Riecher-Rössler A. Prediction of transition to psychosis in patients with a clinical high risk for psychosis: a systematic review of methodology and reporting. Psychological Medicine. 2017;47:1163–1178.

22. Steyerberg EW, Moons KG, van der Windt DA, Hayden JA, Perel P, Schroter S, et al. Prognosis Research Strategy (PROGRESS) 3: prognostic model research. PLoS Medicine. 2013;10:e1001381.

23. Snell KIE, Levis B, Damen JAA, Dhiman P, Debray TPA, Hooft L, et al. Transparent reporting of multivariable prediction models for individual prognosis or diagnosis: checklist for systematic reviews and meta-analyses (TRIPOD-SRMA). BMJ. 2023;381:e073538.

24. Yung AR, Phillips LJ, McGorry PD, McFarlane CA, Francey S, Harrigan S, et al. Prediction of psychosis: a step towards indicated prevention of schizophrenia. The British Journal of Psychiatry. 1998;172:14–20.

25. Geersing G-J, Bouwmeester W, Zuithoff P, Spijker R, Leeflang M, Moons K. Search filters for finding prognostic and diagnostic prediction studies in Medline to enhance systematic reviews. PloS One. 2012;7:e32844.

26. Bramer WM, Giustini D, de Jonge GB, Holland L, Bekhuis T. De-duplication of database search results for systematic reviews in EndNote. Journal of the Medical Library Association: JMLA. 2016;104:240.

27. Moons KG, de Groot JA, Bouwmeester W, Vergouwe Y, Mallett S, Altman DG, et al. Critical appraisal and data extraction for systematic reviews of prediction modelling studies: the CHARMS checklist. PLoS Medicine. 2014;11:e1001744.

28. Harrell Jr FE. Regression Modeling Strategies With Applications to Linear Models, Logistic and Ordinal Regression, and Survival Analysis. 2nd ed. Springer Cham; 2015.

29. Van Calster B, McLernon DJ, van Smeden M, Wynants L, Steyerberg EW, Bossuyt P, et al. Calibration: the Achilles heel of predictive analytics. BMC Medicine. 2019;17:230.

30. Wolff RF, Moons KGM, Riley RD, Whiting PF, Westwood M, Collins GS, et al. PROBAST: A Tool to Assess the Risk of Bias and Applicability of Prediction Model Studies. Ann Intern Med. 2019;170:51–58.

31. Moons KG, Wolff RF, Riley RD, Whiting PF, Westwood M, Collins GS, et al. PROBAST: a tool to assess risk of bias and applicability of prediction model studies: explanation and elaboration. Annals of Internal Medicine. 2019;170:W1–W33.

32. McGuinness LA, Higgins JPT. Risk-of-bias VISualization (robvis): An R package and Shiny web app for visualizing risk-of-bias assessments. Research Synthesis Methods. 2021;12:55–61.

33. Pavlou M, Qu C, Omar RZ, Seaman SR, Steyerberg EW, White IR, et al. Estimation of required sample size for external validation of risk models for binary outcomes. Stat Methods Med Res. 2021;30:2187–2206.

34. Popay J, Roberts H, Sowden A, Petticrew M, Arai L, Rodgers M, et al. Guidance on the conduct of narrative synthesis in systematic reviews. A Product from the ESRC Methods Programme Version. 2006;1:b92.

35. Debray TP, Damen JA, Riley RD, Snell K, Reitsma JB, Hooft L, et al. A framework for meta-analysis of prediction model studies with binary and time-to-event outcomes. Statistical Methods in Medical Research. 2019;28:2768–2786.

36. Perkins DO, Jeffries CD, Addington J, Bearden CE, Cadenhead KS, Cannon TD, et al. Towards a psychosis risk blood diagnostic for persons experiencing high-risk symptoms: preliminary results from the NAPLS project. Schizophr Bull. 2015;41:419–428.

37. Clark S, Baune B, Schubert K, Lavoie S, Smesny S, Rice S, et al. Prediction of transition from ultra-high risk to first-episode psychosis using a probabilistic model combining history, clinical assessment and fatty-acid biomarkers. Translational Psychiatry. 2016;6:e897–e897.

38. Jeffries C, Perkins D, Chandler S, Stark T, Yeo E, Addington J, et al. Insights into psychosis risk from leukocyte microRNA expression. Translational Psychiatry. 2016;6:e981–e981.

39. Lavoie S, Berger M, Schlögelhofer M, Schäfer M, Rice S, Kim S, et al. Erythrocyte glutathione levels as long-term predictor of transition to psychosis. Translational Psychiatry. 2017;7:e1064– e1064.

40. Bousman CA, Lee TY, Kim M, Lee J, Mostaid MS, Bang M, et al. Genetic variation in cytokine genes and risk for transition to psychosis among individuals at ultra-high risk. Schizophrenia Research. 2018;195:589–590.

41. Perkins DO, Olde Loohuis L, Barbee J, Ford J, Jeffries CD, Addington J, et al. Polygenic risk score contribution to psychosis prediction in a target population of persons at clinical high risk. American Journal of Psychiatry. 2020;177:155–163.

42. Mondelli V, Blackman G, Kempton MJ, Pollak TA, Iyegbe C, Valmaggia LR, et al. Serum immune markers and transition to psychosis in individuals at clinical high risk. Brain, Behavior, and Immunity. 2023;110:290–296.

43. Tavares V, Vassos E, Marquand A, Stone J, Valli I, Barker GJ, et al. Prediction of transition to psychosis from an at-risk mental state using structural neuroimaging, genetic, and environmental data. Frontiers in Psychiatry. 2022;13.

44. Zhang T, Zeng J, Wei Y, Ye J, Tang X, Xu L, et al. Changes in inflammatory balance correlates with conversion to psychosis among individuals at clinical high-risk: A prospective cohort study. Psychiatry Research. 2022;318:114938.

45. Zhang T, Wei Y, Zeng J, Ye J, Tang X, Xu L, et al. Interleukin-2/interleukin-6 imbalance correlates with conversion to psychosis from a clinical high-risk state. Psychiatry and Clinical Neurosciences. 2023;77:62–63.

46. Zhang T, Zeng J, Ye J, Gao Y, Hu Y, Xu L, et al. Serum complement proteins rather than inflammatory factors is effective in predicting psychosis in individuals at clinical high risk. Translational Psychiatry. 2023;13:9.

47. Chan MK, Krebs M, Cox D, Guest P, Yolken R, Rahmoune H, et al. Development of a blood-based molecular biomarker test for identification of schizophrenia before disease onset. Translational Psychiatry. 2015;5:e601–e601.

48. Collins GS, Ogundimu EO, Altman DG. Sample size considerations for the external validation of a multivariable prognostic model: a resampling study. Statistics in Medicine. 2016;35:214–226.

49. Riley RD, Debray TPA, Collins GS, Archer L, Ensor J, van Smeden M, et al. Minimum sample size for external validation of a clinical prediction model with a binary outcome. Statistics in Medicine. 2021;40:4230–4251.

50. Damen JAA, Moons KGM, van Smeden M, Hooft L. How to conduct a systematic review and meta-analysis of prognostic model studies. Clinical Microbiology and Infection. 10.1016/j.cmi.2022.07.019.

51. Moriarty AS, Meader N, Snell KI, Riley RD, Paton LW, Dawson S, et al. Predicting relapse or recurrence of depression: systematic review of prognostic models. The British Journal of Psychiatry. 2022:1–11.

52. Lee R, Leighton SP, Thomas L, Gkoutos GV, Wood SJ, Fenton S-JH, et al. Prediction models in first-episode psychosis: systematic review and critical appraisal. The British Journal of Psychiatry. 2022:1–13.

53. Wallace E, Smith SM, Perera-Salazar R, Vaucher P, McCowan C, Collins G, et al. Framework for the impact analysis and implementation of Clinical Prediction Rules (CPRs). BMC Medical Informatics and Decision Making. 2011;11:62.

54. Collins GS, Reitsma JB, Altman DG, Moons KG. Transparent reporting of a multivariable prediction model for individual prognosis or diagnosis (TRIPOD): the TRIPOD statement. Journal of British Surgery. 2015;102:148–158.

55. Burger B, Vaudel M, Barsnes H. Importance of block randomization when designing proteomics experiments. Journal of Proteome Research. 2020;20:122–128.

56. Qin L-X, Zhou Q, Bogomolniy F, Villafania L, Olvera N, Cavatore M, et al. Blocking and Randomization to Improve Molecular Biomarker DiscoveryBlocking and Randomization for Biomarker Discovery. Clinical Cancer Research. 2014;20:3371–3378.

57. Lång U, Yates K, Leacy FP, Clarke MC, McNicholas F, Cannon M, et al. Systematic Review and Meta-analysis: Psychosis Risk in Children and Adolescents With an At-Risk Mental State. Journal of the American Academy of Child & Adolescent Psychiatry. 2022;61:615–625.

58. Ajnakina O, Morgan C, Gayer-Anderson C, Oduola S, Bourque F, Bramley S, et al. Only a small proportion of patients with first episode psychosis come via prodromal services: a retrospective survey of a large UK mental health programme. BMC Psychiatry. 2017;17:1–9.

59. Burke T, Thompson A, Mifsud N, Yung AR, Nelson B, McGorry P, et al. Proportion and characteristics of young people in a first-episode psychosis clinic who first attended an at-risk mental state service or other specialist youth mental health service. Schizophrenia Research. 2022;241:94–101.

60. Bolhuis K, Lång U, Gyllenberg D, Kääriälä A, Veijola J, Gissler M, et al. Hospital presentation for self-harm in youth as a risk marker for later psychotic and bipolar disorders: a cohort study of 59 476 Finns. Schizophrenia Bulletin. 2021;47:1685–1694.

61. Lång U, Ramsay H, Yates K, Veijola J, Gyllenberg D, Clarke MC, et al. Potential for prediction of psychosis and bipolar disorder in Child and Adolescent Mental Health Services: a longitudinal register study of all people born in Finland in 1987. World Psychiatry. 2022;21:436–443.

62. Cotter D, Healy C, Staines L, Mongan D, Cannon M. Broadening the parameters of clinical high risk for psychosis. American Journal of Psychiatry. 2022;179:593–595.

63. Wigman JT, van Nierop M, Vollebergh WA, Lieb R, Beesdo-Baum K, Wittchen H-U, et al. Evidence that psychotic symptoms are prevalent in disorders of anxiety and depression, impacting on illness onset, risk, and severity—implications for diagnosis and ultra–high risk research. Schizophrenia Bulletin. 2012;38:247–257.

64. Fusar-Poli P, Rutigliano G, Stahl D, Davies C, Bonoldi I, Reilly T, et al. Development and Validation of a Clinically Based Risk Calculator for the Transdiagnostic Prediction of Psychosis. JAMA Psychiatry. 2017;74:493–500.

65. Fusar-Poli P, Werbeloff N, Rutigliano G, Oliver D, Davies C, Stahl D, et al. Transdiagnostic Risk Calculator for the Automatic Detection of Individuals at Risk and the Prediction of Psychosis: Second Replication in an Independent National Health Service Trust. Schizophrenia Bulletin. 2019;45:562–570.

66. Oliver D, Wong CMJ, Bøg M, Jönsson L, Kinon BJ, Wehnert A, et al. Transdiagnostic individualized clinically-based risk calculator for the automatic detection of individuals at-risk and the prediction of psychosis: external replication in 2,430,333 US patients. Translational Psychiatry. 2020;10:364.

67. Martin B, Wittmann M, Franck N, Cermolacce M, Berna F, Giersch A. Temporal structure of consciousness and minimal self in schizophrenia. Frontiers in Psychology. 2014;5.

68. Lunsford-Avery JR, Gonçalves B da SB, Brietzke E, Bressan RA, Gadelha A, Auerbach RP, et al. Adolescents at clinical-high risk for psychosis: Circadian rhythm disturbances predict worsened prognosis at 1-year follow-up. Schizophrenia Research. 2017;189:37–42.

69. Clementz BA, Parker DA, Trotti RL, McDowell JE, Keedy SK, Keshavan MS, et al. Psychosis Biotypes: Replication and Validation from the B-SNIP Consortium. Schizophrenia Bulletin. 2022;48:56–68.

70. Sun G-W, Shook TL, Kay GL. Inappropriate use of bivariable analysis to screen risk factors for use in multivariable analysis. Journal of Clinical Epidemiology. 1996;49:907–916.

71. Riley RD, Snell KIE, Martin GP, Whittle R, Archer L, Sperrin M, et al. Penalization and shrinkage methods produced unreliable clinical prediction models especially when sample size was small. Journal of Clinical Epidemiology. 2021;132:88–96.

72. Riley RD, Ensor J, Snell KI, Harrell FE, Martin GP, Reitsma JB, et al. Calculating the sample size required for developing a clinical prediction model. Bmj. 2020;368.

73. Riley RD, Snell KI, Ensor J, Burke DL, Harrell Jr FE, Moons KG, et al. Minimum sample size for developing a multivariable prediction model: PART II - binary and time-to-event outcomes. Statistics in Medicine. 2019;38:1276–1296.

74. Vickers AJ, van Calster B, Steyerberg EW. A simple, step-by-step guide to interpreting decision curve analysis. Diagnostic and Prognostic Research. 2019;3:18.

75. Gardner ML, Freitas MA. Multiple Imputation Approaches Applied to the Missing Value Problem in Bottom-Up Proteomics. International Journal of Molecular Sciences. 2021;22.

76. van Os J, Guloksuz S. A critique of the “ultra-high risk” and “transition” paradigm. World Psychiatry. 2017;16:200–206.

