## Supplementary Materials for "Prognostic models predicting transition to psychotic disorder using blood-based biomarkers: a systematic review and critical appraisal"

### Supplementary Methods

### Supplementary Table 1: TRIPOD-SRMA

| Section and topic | Item No | Checklist item | Page |
|---|---|---|---|
| **Title** | | | |
| Title | 1 | Identify the report as a systematic review or meta-analysis (or both) of diagnostic or prognostic model studies. Specify the target population and outcome(s) predicted as relevant to the review question. | Title (p1) |
| **Abstract** | | | |
| Abstract | 2 | See the TRIPOD-SRMA Checklist for Abstracts | Abstract (p2) |
| **Introduction** | | | |
| Rationale | 3 | Describe the rationale for the review in the context of existing knowledge. | Intro. (p3) |
| Objectives | 4 | Provide an explicit statement of the objective(s) being addressed with reference to: target population, index and comparator models (as relevant), outcome(s), time (prediction horizon and intended moment of using the model), and setting. | Intro. (p4), s. Table 2 |
| **Methods** | | | |
| Study eligibility criteria | 5 | Specify study characteristics used as eligibility criteria, including any prediction models of specific interest, and whether development or validation studies (or both) were eligible. | Methods (p4) |
| Information sources | 6 | Specify all databases, registers, websites, organisations, reference lists and other sources searched or consulted to identify studies. Specify the date when each source was last searched or consulted. | Methods (p5-6) |
| Search strategy | 7 | Present the full search strategies for all databases, registers and websites, including any filters and limits used. | Supp. (p5-8) |
| Study selection process | 8 | Specify the methods used to decide whether a study met the inclusion criteria of the review, including how many reviewers screened each record and each report retrieved, whether they worked independently, and if applicable, details of automation tools used in the process. | Methods (p6) |
| Data collection process | 9 | Specify methods used to collect data from study reports, including how many reviewers collected data from each report, whether they worked independently, any processes for obtaining or confirming data from study investigators, and if applicable, details of automation tools used in the process. | Methods (p6-7) |
| Data Items | 10a | List and define all items for which data were sought from each study. | Methods (p6-7) |
| | 10b | State the model performance measures that were sought (e.g., measures of calibration, discrimination, overall model fit, clinical utility). | Methods (p7) |
| | 10c | Describe how any desired but unreported data items (items 10a, 10b) were handled (e.g., contacted authors, calculated from other reported information). | Methods (p6-7) |
| Risk of bias and applicability assessment | 11 | Specify the methods used to assess risk of bias in the included studies and their applicability to the review question. This should be done separately for each model development and validation. Include details of any tool(s) used, how many reviewers assessed each study and whether they worked independently. | Methods (p7) |
| Synthesis methods | 12a | Describe any methods for synthesising estimates of performance measures for each model. If meta-analysis was carried out, describe the methods used, including any transformations of data prior to pooling, how any heterogeneity in model performance was quantified and handled, and software package(s) used. | Methods (p7-8) |
| | 12b | Describe any methods used to explore possible causes of heterogeneity in model performance (e.g., subgroup analysis, meta-regression), including whether or not they were planned. | N/A |

| Section and topic | Item No | Checklist item | Page |
|---|---|---|---|
| | 12c | Describe any sensitivity analyses conducted to assess robustness of the synthesised results. | Methods (p7-8) |
| Certainty assessment | 13 | Describe any methods used to assess certainty (or confidence) in the body of evidence for a prediction model. | N/A |
| **Results** | | | |
| Study selection | 14 | Describe the results of the search and selection process, from the number of records identified in the search to the number of studies and models included in the review, ideally using a flow diagram. | Results (p8) |
| Study and model characteristics | 15 | Present study characteristics and model details extracted (as per Item 10a), and cite the study reports. | Results (p9-11), Table 1, Supp. (p9-23) |
| Risk of bias and applicability | 16 | Present results of risk of bias and applicability assessment. This should be done separately for each model development and validation in each included study. | Results (p10-11) |
| Results of model performance in individual studies | 17 | Present performance estimates and confidence intervals for each model and all evaluations, including whether they relate to the internal or external validation performance. If internal, give details of the method. | Results (p11-12), Table 1, Supp. (p9-17) |
| Results of syntheses | 18a | Present the results of any synthesis of model performance, together with details of which study estimates contributed. If meta-analysis was carried out, then for each model and performance measure, present summary results, confidence/credible intervals and measures of heterogeneity. Forest plots may be useful. | Results (p11-12) |
| | 18b | For each model, present results of all investigations of possible causes of heterogeneity in model performance. | N/A |
| | 18c | Present results of all sensitivity analyses conducted to assess the robustness of the synthesised results. | Results (p11-12) |
| Certainty of evidence | 19 | Present any assessments of certainty (or confidence) in the body of evidence for each prediction model of interest. | N/A |
| **Discussion** | | | |
| Summary of evidence | 20 | Summarise the main findings including the strengths and limitations of the evidence. | Discussion (p12-16) |
| Limitations | 21 | Discuss the strengths and limitations of the review process. | Discussion (p16) |
| Implications | 22 | Discuss implications of the results in the context of other evidence and for practice, policy, and future research. | Discussion (p12-16) |
| **Other information** | | | |
| Registration and protocol | 23a | Provide registration information for the review, including register name and registration number, or state that the review was not registered. | Registration (p17) |
| | 23b | Indicate where the review protocol can be accessed, or state that a protocol was not prepared. | (p17) |
| | 23c | Describe and explain any amendments to information provided at registration or in the protocol. | N/A |
| Support | 24 | Describe sources of financial or non-financial support for the review, and the role of the funders or sponsors in the review. | Funding (p17) |
| Competing interests | 25 | Declare any competing interests of review authors. | Conflict of Interest (p17) |

| Section and topic | Item No | Checklist item | Page |
|---|---|---|---|
| Availability of data, code, and other materials | 26 | Report which of the following are publicly available and where they can be found: template data collection forms; data extracted from included studies; data used for all analyses; analytic code; any other materials used in the review. | Methods (p6-7), Table 1, Supp. (p9-23) |

This checklist appears in appendix 2 of Snell KIE, Levis B, Damen JAA, et al. Transparent reporting of multivariable prediction models for individual prognosisor diagnosis: checklist for systematic reviews and meta-analyses (TRIPOD-SRMA). *BMJ* 2023;381:e073538. doi:10.1136/bmj-2022-073538.

#### Supplementary Table 2: PICOTS Criteria

| | |
|---|---|
| **Participants** | Clinical high-risk for psychosis, at-risk mental state or similar enrichment for psychosis risk. |
| **Intervention/Index** | Prognostic models using blood-biomarkers. Model development, validation or updating. |
| **Comparator** | Prognostic models using predictors other than blood-biomarkers. |
| **Outcome** | Transition to psychosis. |
| **Timing** | Models predicting transition at any time before onset to a ten-year horizon. |
| **Setting** | Any setting. To support early intervention strategies and trials of preventative therapies in individuals at risk of psychosis. |

### Search Strategy

#### Pubmed

"clinical high-risk"[Title/Abstract] OR "clinically high-risk"[Title/Abstract] OR "ultra high-risk"[Title/Abstract] OR "at-risk mental state"[Title/Abstract] OR "prodrom*"[Title/Abstract] OR "self-harm"[Title/Abstract] OR "risk of psychosis"[Title/Abstract] OR "attenuated psychotic symptoms"[Title/Abstract] OR "brief and limited intermittent psychotic symptoms"[Title/Abstract] OR "genetic risk and deterioration syndrome"[Title/Abstract] OR "psychotic experiences"[Title/Abstract] OR "help-seeking"[Title/Abstract] OR "psychosis-like symptoms"[Title/Abstract] OR "psychosis risk syndrome"[Title/Abstract] OR "familial high risk"[Title/Abstract] OR "familial risk"[Title/Abstract] OR "mental health services"[MeSH Terms] OR (("psychotic symptoms"[Title/Abstract] OR "psychosis"[Title/Abstract] OR "psychotic disorder"[Title/Abstract] OR "schiz*"[Title/Abstract] OR "bipolar"[Title/Abstract] OR "delusion*"[Title/Abstract] OR "schizophrenia spectrum and other psychotic disorders"[MeSH Terms]) AND ("sub-threshold"[Title/Abstract] OR "sub-clinical"[Title/Abstract] OR "non-clinical"[Title/Abstract]))

AND ("psychosis"[Title/Abstract] OR "psychotic disorder"[Title/Abstract] OR "schiz*"[Title/Abstract] OR "bipolar disorder"[Title/Abstract] OR "delusion*"[Title/Abstract] OR "schizophrenia spectrum and other psychotic disorders"[MeSH Terms]) AND ("conver*"[Title/Abstract] OR "transition*"[Title/Abstract] OR "develop*"[Title/Abstract] OR "later*"[Title/Abstract] OR "follow up"[Title/Abstract] OR "follow up"[Title/Abstract] OR "onset"[Title/Abstract] OR "progress*"[Title/Abstract] OR "prior"[Title/Abstract] OR "before"[Title/Abstract])

AND "prote*"[Title/Abstract] OR "nucleic acid"[Title/Abstract] OR "RNA"[Title/Abstract] OR "mRNA"[Title/Abstract] OR "miRNA"[Title/Abstract] OR "microRNA"[Title/Abstract] OR "DNA"[Title/Abstract] OR "genetic"[Title/Abstract] OR "genom*"[Title/Abstract] OR "epigen*"[Title/Abstract] OR "cytokine*"[Title/Abstract] OR "chemokine*"[Title/Abstract] OR "inflammatory"[Title/Abstract] OR "immune"[Title/Abstract] OR "lipid*"[Title/Abstract] OR "apolipo*"[Title/Abstract] OR "phospholipid*"[Title/Abstract] OR "autoantibod*"[Title/Abstract] OR "neurotransmitter"[Title/Abstract] OR "blood"[Title/Abstract] OR "plasma"[Title/Abstract] OR "serum"[Title/Abstract] OR "peripheral biomarkers"[Title/Abstract] OR "biomarkers/blood"[MeSH Terms] OR "amino acids, peptides, and proteins"[MeSH Terms] OR "nucleic acids, nucleotides, and nucleosides"[MeSH Terms] OR "Lipids"[MeSH Terms]

AND (("validat*"[Title/Abstract] OR "predict*"[Title/Abstract] OR "rule*"[Title/Abstract]) OR ("predict*"[Title/Abstract] AND ("outcome*"[Title/Abstract] OR "risk*"[Title/Abstract] OR "model*"[Title/Abstract])) OR (("History"[Title/Abstract] OR "variable*"[Title/Abstract] OR "Criteria"[Title/Abstract] OR "scor*"[Title/Abstract] OR "characteristic*"[Title/Abstract] OR "finding*"[Title/Abstract] OR "factor*"[Title/Abstract]) AND ("predict*"[Title/Abstract] OR "model*"[Title/Abstract] OR "decision*"[Title/Abstract] OR "identif*"[Title/Abstract] OR "prognos*"[Title/Abstract])) OR ("decision*"[Title/Abstract] AND ("model*"[Title/Abstract] OR "clinical*"[Title/Abstract] OR "logistic models"[Title/Abstract])) OR ("Prognostic"[Title/Abstract] AND ("History"[Title/Abstract] OR "variable*"[Title/Abstract] OR "Criteria"[Title/Abstract] OR "scor*"[Title/Abstract] OR "characteristic*"[Title/Abstract] OR "finding*"[Title/Abstract] OR "factor*"[Title/Abstract] OR "model*"[Title/Abstract])) OR ("stratification"[Title/Abstract] OR "roc curve"[MeSH Terms] OR "discrimination"[Title/Abstract] OR "discriminate"[Title/Abstract] OR "c statistic"[Title/Abstract] OR "c statistic"[Title/Abstract] OR "area under the curve"[Title/Abstract] OR "auc"[Title/Abstract] OR "calibration"[Title/Abstract] OR "indices"[Title/Abstract] OR "algorithm"[Title/Abstract] OR "multivariable"[Title/Abstract] OR "machine learning"[Title/Abstract] OR "artificial intelligence"[Title/Abstract] OR "deep learning"[Title/Abstract] OR "algorithm"[Title/Abstract]))

AND 1998/01/01:2023/05/15[Date - Publication]

**EMBASE**

'clinical high-risk':ti,ab OR 'clinically high-risk':ti,ab OR 'ultra high-risk':ti,ab OR 'at-risk mental state':ti,ab OR 'prodrom*':ti,ab OR 'self-harm':ti,ab OR 'risk of psychosis':ti,ab OR 'attenuated psychotic symptoms':ti,ab OR 'brief and limited intermittent psychotic symptoms':ti,ab OR 'genetic risk and deterioration syndrome':ti,ab OR 'psychotic experiences':ti,ab OR 'help-seeking':ti,ab OR 'psychosis-like symptoms':ti,ab OR 'psychosis risk syndrome':ti,ab OR 'familial high risk':ti,ab OR 'familial risk':ti,ab OR ‘psychiatric emergency service’/exp OR ‘mental health service’/exp OR (('psychotic symptoms':ti,ab OR 'psychosis':ti,ab OR 'psychotic disorder':ti,ab OR 'schiz*':ti,ab OR 'bipolar':ti,ab OR 'delusion*':ti,ab OR ‘schizophrenia spectrum disorder’/exp) AND ('sub-threshold':ti,ab OR 'sub-clinical':ti,ab OR 'non-clinical':ti,ab))

AND (("psychosis":ti,ab OR "psychotic disorder":ti,ab OR "schiz*":ti,ab OR "bipolar disorder":ti,ab OR "delusion*":ti,ab OR ‘schizophrenia spectrum disorder’/exp) AND (conver*:ti,ab OR transition*:ti,ab OR develop*:ti,ab OR later*:ti,ab OR 'follow up':ti,ab OR 'follow-up':ti,ab OR onset OR ‘progress*’:ti,ab OR ‘prior’:ti,ab OR ‘before’:ti,ab))

AND ('prote*':ti,ab OR 'nucleic acid':ti,ab OR 'rna':ti,ab OR 'mrna':ti,ab OR 'mirna':ti,ab OR 'microrna':ti,ab OR 'dna':ti,ab OR 'genetic':ti,ab OR 'genome':ti,ab OR 'epigen*':ti,ab OR 'cytokine*':ti,ab OR 'chemokine*':ti,ab OR 'inflammatory':ti,ab OR ‘immune’:ti,ab OR 'lipid*':ti,ab OR 'fatty acid*':ti,ab OR 'apolipo*':ti,ab OR 'phospholipid*':ti,ab OR 'autoantibod*':ti,ab OR 'neurotransmitter':ti,ab OR 'blood':ti,ab OR 'plasma':ti,ab OR 'serum':ti,ab OR 'peripheral biomarkers':ti,ab OR 'peptides and proteins'/exp OR 'nucleic acid'/exp OR 'nucleoside'/exp OR 'nucleotide'/exp OR 'steroid'/exp OR 'lipid'/exp OR 'amino acid'/exp)

AND (("validat*":ti,ab OR "predict*":ti,ab OR "rule*":ti,ab) OR ("predict*":ti,ab AND ("outcome*":ti,ab OR "risk*":ti,ab OR "model*":ti,ab)) OR (("History":ti,ab OR "variable*":ti,ab OR "Criteria":ti,ab OR "scor*":ti,ab OR "characteristic*":ti,ab OR "finding*":ti,ab OR "factor*":ti,ab) AND ("predict*":ti,ab OR "model*":ti,ab OR "decision*":ti,ab OR "identif*":ti,ab OR "prognos*":ti,ab)) OR ("decision*":ti,ab AND ("model*":ti,ab OR "clinical*":ti,ab OR "logistic models":ti,ab)) OR ("Prognostic":ti,ab AND ("History":ti,ab OR "variable*":ti,ab OR "Criteria":ti,ab OR "scor*":ti,ab OR "characteristic*":ti,ab OR "finding*":ti,ab OR "factor*":ti,ab OR "model*":ti,ab)) OR ("stratification":ti,ab OR "receiver operating characteristic":ti,ab OR "discrimination":ti,ab OR "discriminate":ti,ab OR "c statistic":ti,ab OR "c statistic":ti,ab OR "area under the curve":ti,ab OR "auc":ti,ab OR "calibration":ti,ab OR "indices":ti,ab OR "algorithm":ti,ab OR "multivariable":ti,ab) OR ("machine learning":ti,ab OR "artificial intelligence":ti,ab OR "deep learning":ti,ab OR "algorithm":ti,ab))

AND ([article]/lim OR [article in press]/lim OR [conference abstract]/lim OR [conference paper]/lim OR [letter]/lim OR [note]/lim) AND ([humans]/lim OR [clinical study]/lim)

AND [1998-2023]/py

**PsycINFO**

(TI "clinical high-risk" OR AB "clinical high-risk") OR (TI "clinically high-risk" OR AB "clinically high-risk") OR (TI "ultra high-risk" OR AB "ultra high-risk") OR (TI "at-risk mental state" OR AB "at-risk mental state") OR (TI prodrom* OR AB prodrom*) OR (TI self-harm OR AB self-harm) OR (TI "risk of psychosis" OR AB "risk of psychosis") OR (TI "attenuated psychotic symptoms" OR AB "attenuated psychotic symptoms") OR (TI "brief and limited intermittent psychotic symptoms" OR AB "brief and limited intermittent psychotic symptoms") OR (TI "genetic risk and deterioration syndrome" OR AB "genetic risk and deterioration syndrome") OR (TI "psychotic experiences" OR AB "psychotic experiences") OR (TI help-seeking OR AB help-seeking) OR (TI "psychosis-like symptoms" OR AB "psychosis-like symptoms") OR (TI "psychosis risk syndrome" OR AB "psychosis risk syndrome") OR (TI "familial high risk" OR AB "familial high risk") OR (TI "familial risk" OR AB "familial risk") OR (((TI "psychotic symptoms" OR AB "psychotic symptoms") OR (TI psychosis OR AB psychosis) OR (TI "psychotic disorder" OR AB "psychotic disorder") OR (TI schiz* OR AB schiz*) OR (TI bipolar OR AB bipolar) OR (TI delusion* OR AB delusion*)) AND ((TI sub-threshold OR AB sub-threshold) OR (TI sub-clinical OR AB sub-clinical) OR (TI non-clinical OR AB non-clinical)))

AND (((TI psychosis OR AB psychosis) OR (TI "psychotic disorder" OR AB "psychotic disorder") OR (TI schiz* OR AB schiz*) OR (TI "bipolar disorder" OR AB "bipolar disorder") OR (TI delusion* OR AB delusion*)) AND ((TI conver* OR AB conver*) OR (TI transition* OR AB transition*) OR (TI develop* OR AB develop*) OR (TI later* OR AB later*) OR (TI "'follow up'" OR AB "'follow up'") OR (TI 'follow-up' OR AB 'follow-up') OR (TI ‘onset’ OR AB ‘onset’) OR (TI progress* OR AB progress*) OR (TI prior OR AB prior) OR (TI before OR AB before)))

AND ((TI prote* OR AB prote*) OR (TI "nucleic acid" OR AB "nucleic acid") OR (TI RNA OR AB RNA) OR (TI mRNA OR AB mRNA) OR (TI miRNA OR AB miRNA) OR (TI microRNA OR AB microRNA) OR (TI DNA OR AB DNA) OR (TI genetic OR AB genetic) OR (TI genome OR AB genome) OR (TI epigen* OR AB epigen*) OR (TI cytokine* OR AB cytokine*) OR (TI chemokine* OR AB chemokine*)OR (TI "inflammatory" OR AB "inflammatory") OR (TI "immune" OR AB "immune") OR (TI lipid* OR AB lipid*) OR (TI apolipo* OR AB apolipo*) OR (TI phospholipid* OR AB phospholipid*) OR (TI autoantibod* OR AB autoantibod*) OR (TI neurotransmitter OR AB neurotransmitter) OR (TI blood OR AB blood) OR (TI plasma OR AB plasma) OR (TI serum OR AB serum) OR (TI "peripheral biomarkers" OR AB "peripheral biomarkers") OR (MH biomarkers/blood+) OR (MH "amino acids, peptides, and proteins"+) OR (MH "nucleic acids, nucleotides, and nucleosides"+) OR (MH Lipids+) OR MA "biomarkers/blood" OR MA ( "amino acids, peptides, and proteins" ) OR MA ( "nucleic acids, nucleotides, and nucleosides" ) OR MA "Lipids") OR (((DE "Amino Acids" OR DE "Alanines" OR DE "Aspartic Acid" OR DE "Beta Amyloid" OR DE "Cysteine" OR DE "DOPA" OR DE "Gamma Aminobutyric Acid" OR DE "Glutamic Acid" OR DE "Glutamine" OR DE "Glycine" OR DE "Histidine" OR DE "Leucine" OR DE "Melanin" OR DE "Methionine" OR DE "Neurokinins" OR DE "Peptides" OR DE "Proline" OR DE "Tryptophan" OR DE "Tyrosine" OR DE "Peptides" OR DE "Angiotensin" OR DE "Bombesin" OR DE "Cholecystokinin" OR DE "Corticotropin Releasing Factor" OR DE "Endogenous Opiates" OR DE "Ghrelin" OR DE "Insulin-like Growth Factor" OR DE "Leptin" OR DE "Melanocyte Stimulating Hormone" OR DE "Nerve Growth Factor" OR DE "Neuropeptides" OR DE "Neurotensin" OR DE "Somatostatin" OR DE "Tachykinins") AND (DE "Neurotransmitters" OR DE "Acetylcholine" OR DE "Aspartic Acid" OR DE "Catecholamines" OR DE "Cholecystokinin" OR DE "Endorphins" OR DE "Gamma Aminobutyric Acid" OR DE "Glutamic Acid" OR DE "Glycine" OR DE "Histamine" OR DE "Neurokinins" OR DE "Neurotensin" OR DE "Serotonin" OR DE "Substance P" OR DE "Proteins" OR DE "Apolipoprotein E" OR DE "Apolipoproteins" OR DE "Beta Amyloid" OR DE "Blood Proteins" OR DE "Cell Adhesion Molecules" OR DE "CLOCK Gene"

OR DE "Endorphins" OR DE "Globulins" OR DE "Growth Factor" OR DE "Interferons" OR DE "Ion Channel" OR DE "Neurofibrillary Tangles" OR DE "Neurotransmitter Transporters" OR DE "Prion" OR DE "Rhodopsin" OR DE "Synaptotagmin" OR DE "Tau Proteins")) OR (DE "Nucleic Acids" OR DE "Adenosine" OR DE "DNA" OR DE "Nucleotides" OR DE "Ribonucleic Acid")) OR (DE "Lipids" OR DE "Fatty Acids" OR DE "Gangliosides" OR DE "Lipopolysaccharide" OR DE "Lipoproteins")

AND (((TI validat* OR AB validat*) OR (TI predict* OR AB predict*) OR (TI rule* OR AB rule*)) OR ((TI predict* OR AB predict*) AND ((TI outcome* OR AB outcome*) OR (TI risk* OR AB risk*) OR (TI model* OR AB model*))) OR (((TI History OR AB History) OR (TI variable* OR AB variable*) OR (TI Criteria OR AB Criteria) OR (TI scor* OR AB scor*) OR (TI characteristic* OR AB characteristic*) OR (TI finding* OR AB finding*) OR (TI factor* OR AB factor*)) AND ((TI predict* OR AB predict*) OR (TI model* OR AB model*) OR (TI decision* OR AB decision*) OR (TI identif* OR AB identif*) OR (TI prognos* OR AB prognos*))) OR ((TI decision* OR AB decision*) AND ((TI model* OR AB model*) OR (TI clinical* OR AB clinical*) OR (TI "logistic models" OR AB "logistic models"))) OR ((TI Prognostic OR AB Prognostic) AND ((TI History OR AB History) OR (TI variable* OR AB variable*) OR (TI Criteria OR AB Criteria) OR (TI scor* OR AB scor*) OR (TI characteristic* OR AB characteristic*) OR (TI finding* OR AB finding*) OR (TI factor* OR AB factor*) OR (TI model* OR AB model*))) OR ((TI stratification OR AB stratification) OR (MH "roc curve"+) OR (TI discrimination OR AB discrimination) OR (TI discriminate OR AB discriminate) OR (TI "c statistic" OR AB "c statistic") OR (TI "c statistic" OR AB "c statistic") OR (TI "area under the curve" OR AB "area under the curve") OR (TI auc OR AB auc) OR (TI calibration OR AB calibration) OR (TI indices OR AB indices) OR (TI algorithm OR AB algorithm) OR (TI multivariable OR AB multivariable) OR (TI "machine learning" OR AB "machine learning") OR (TI "artificial intelligence" OR AB "artificial intelligence") OR (TI "deep learning" OR AB "deep learning") OR (TI algorithm OR AB algorithm)))

### Supplementary Results

#### Study Characteristics

| | |
|---|---|
| **Study** | Clark et al., 2016 |
| **Transition definition and method of diagnosis** | PANSS Scale cut-offs: score of ≥4 on hallucinations, ≥4 on delusions, or ≥5 on conceptual disorganization). These levels had to be sustained for at least 1 week. |
| **Time of outcome occurrence or summary of duration of follow-up:** | 1 year |
| **Method for selection of predictors for inclusion in multivariable modelling** | Univariate AUROC. |
| **Method for selection of predictors during multivariable modelling** | Forward selection. |
| **Shrinkage of predictor weights or regression coefficients** | None |
| **Discrimination (95% CI)** | C-index 0.92 (0.786–0.982) |
| **Calibration** | NI |

| | |
|---|---|
| **Study** | Dickens et al., 2021 |
| **Transition definition and method of diagnosis** | At follow-up, trained raters<br>assessed participants using the CAARMS |
| **Time of outcome occurrence or summary of duration of follow-up:** | Up to 5 years. |
| **Method for selection of predictors for inclusion in multivariable modelling** | Pre-selection based on univariate testing ($p < 0.5$) and multivariate analyses. |
| **Method for selection of predictors during multivariable modelling** | Backwards selection. |
| **Shrinkage of predictor weights or regression coefficients** | L2 regularisation. |
| **Discrimination (95% CI)** | C-index 0.81 (0.69–0.93) |
| **Calibration** | NI |

| | |
|---|---|
| **Study** | Chan et al., 2015 |
| **Transition definition and method of diagnosis** | Psychosis was defined by a score of >6 on the CAARMS sub-scale of thought disorder, >5 on the perceptual abnormalities sub-scale and/or >6 on the disorganized speech sub-scale with frequency scores ≥4 on the thought disorder, perceptual abnormalities and/or disorganized speech sub-scales for more than one week. |
| **Time of outcome occurrence or summary of duration of follow-up:** | Unclear |
| **Method for selection of predictors for inclusion in multivariable modelling** | Based on previous discovery case-control cohorts. |
| **Method for selection of predictors during multivariable modelling** | L1 regularisation. |
| **Shrinkage of predictor weights or regression coefficients** | L1 regularisation. |
| **Discrimination (95% CI)** | C-index 0.90 (0.82-0.87) |
| **Calibration** | NI |

| | |
|---|---|
| **Study** | Jeffries et al., 2016 |
| **Transition definition and method of diagnosis** | The Structured Clinical Interview for DSM-IV was used to determine psychiatric diagnoses. |
| **Time of outcome occurrence or summary of duration of follow-up:** | Up to 2 years |
| **Method for selection of predictors for inclusion in multivariable modelling** | All candidate predictors with reads in 90% of subjects were included. |
| **Method for selection of predictors during multivariable modelling** | Similar to forward selection, predictors were added that maximised the P-value further. |
| **Shrinkage of predictor weights or regression coefficients** | Vector (+/-1) coefficients |
| **Discrimination (95% CI)** | C-index 0.86 (NI) |
| **Calibration** | NI |

| | |
|---|---|
| **Study** | Koutsouleris et al., 2021 |
| **Transition definition and method of diagnosis** | At least 1 of the 5 positive symptom items in the Structured Interview for Psychosis–Risk Syndromes reached psychotic intensity daily for at least 7 days. |
| **Time of outcome occurrence or summary of duration of follow-up:** | 18-36 months. |
| **Method for selection of predictors for inclusion in multivariable modelling** | Data reduction methods. |
| **Method for selection of predictors during multivariable modelling** | Greedy sequential backwards elimination. |
| **Shrinkage of predictor weights or regression coefficients** | L2 regularization |
| **Discrimination (95% CI)** | C-index 0.74 (NI); BAC 66.1% (NI) |
| **Calibration** | NI |

| | |
|---|---|
| **Study** | Lavoie et al., 2017 |
| **Transition definition and method of diagnosis** | PANSS Scale cut-offs: score of ≥4 on hallucinations, ≥4 on delusions, or ≥5 on conceptual disorganization). These levels had to be sustained for at least 1 week. |
| **Time of outcome occurrence or summary of duration of follow-up:** | 7 years |
| **Method for selection of predictors for inclusion in multivariable modelling** | Unclear. |
| **Method for selection of predictors during multivariable modelling** | N/A |
| **Shrinkage of predictor weights or regression coefficients** | None |
| **Discrimination (95% CI)** | C-index 0.82 (NI) |
| **Calibration** | NI |

| | |
|---|---|
| **Study** | Bousman et al., 2018 |
| **Transition definition and method of diagnosis** | At least one<br>fully positive psychotic symptom several times a week for over one week as measured by the SIPS |
| **Time of outcome occurrence or summary of duration of follow-up:** | Up to 7.7 years |
| **Method for selection of predictors for inclusion in multivariable modelling** | Univariate screening. |
| **Method for selection of predictors during multivariable modelling** | N/A |
| **Shrinkage of predictor weights or regression coefficients** | N/A |
| **Discrimination (95% CI)** | Accuracy 65% (53%-75%) |
| **Calibration** | NI |

| | |
|---|---|
| **Study** | Li et al., 2022 |
| **Transition definition and method of diagnosis** | According<br>to the criteria of Presence of Psychotic Symptoms (POPS) in SIPS/SOPS. |
| **Time of outcome occurrence or summary of duration of follow-up:** | At least 2 years. |
| **Method for selection of predictors for inclusion in multivariable modelling** | Univariate screening. |
| **Method for selection of predictors during multivariable modelling** | N/A |
| **Shrinkage of predictor weights or regression coefficients** | No shrinkage/NI |
| **Discrimination (95% CI)** | C-index 1 (NI) |
| **Calibration** | NI |

| | |
|---|---|
| **Study** | Mongan et al., 2021 |
| **Transition definition and method of diagnosis** | The onset of nonorganic psychotic disorder as assessed either by CAARMS interview or by contact with the clinical team or review of clinical records. |
| **Time of outcome occurrence or summary of duration of follow-up:** | At least 2 years |
| **Method for selection of predictors for inclusion in multivariable modelling** | Proteomic predictors with <20% missing values, otherwise all candidate predictors. |
| **Method for selection of predictors during multivariable modelling** | None (Top ten highest weighted predictors in separate model). |
| **Shrinkage of predictor weights or regression coefficients** | L2 regularisation. |
| **Discrimination (95% CI)** | C-index 0.95 (0.91-0.99) |
| **Calibration** | NI |

| | |
|---|---|
| **Study** | Mondelli et al., 2023 |
| **Transition definition and method of diagnosis** | Transition to psychosis was defined as the development of full threshold psychotic disorder according to CAARMS. |
| **Time of outcome occurrence or summary of duration of follow-up:** | 2 years |
| **Method for selection of predictors for inclusion in multivariable modelling** | Markers with <20% missing values, otherwise all candidate predictors. |
| **Method for selection of predictors during multivariable modelling** | Elastic Net |
| **Shrinkage of predictor weights or regression coefficients** | Elastic Net |
| **Discrimination (95% CI)** | C-index 0.57 (0.52-0.61) |
| **Calibration** | NI |

| | |
|---|---|
| **Study** | Perkins et al., 2015 |
| **Transition definition and method of diagnosis** | Psychosis criteria were met if the subject had a score of “6” on any of the SOPS positive items occurring for at least one hour per day at least four days in a month, with the symptoms seriously disorganizing or dangerous. |
| **Time of outcome occurrence or summary of duration of follow-up:** | Up to 2 years |
| **Method for selection of predictors for inclusion in multivariable modelling** | Analytes that were not detected in ≥ 20% of the subjects were excluded. Analytes with a possible relation to prescription of antipsychotics or antidepressants, or with self-reported current use of marijuana, nicotine, or alcohol, based on comparisons of analyte expression were excluded. |
| **Method for selection of predictors during multivariable modelling** | Similar to forward selection, predictors were added that maximised the P-value further. Top 15 most frequently chosen analytes in bootstraps were retained. |
| **Shrinkage of predictor weights or regression coefficients** | None |
| **Discrimination (95% CI)** | C-index 0.88 (SE 0.043) |
| **Calibration** | NI |

| | |
|---|---|
| **Study** | Perkins et al., 2020 |
| **Transition definition and method of diagnosis** | Psychosis conversion was defined by the “presence of psychosis” criteria (30, 31) (psychotic-severity positive symptoms that are seriously disorganizing or dangerous, and occur at least 1 hour/ day on average 4 days a week) |
| **Time of outcome occurrence or summary of duration of follow-up:** | 2 years |
| **Method for selection of predictors for inclusion in multivariable modelling** | Predictors specified a priori. |
| **Method for selection of predictors during multivariable modelling** | N/A |
| **Shrinkage of predictor weights or regression coefficients** | Unclear |
| **Discrimination (95% CI)** | C-index 0.67 (non-Europeans); 0.71 (Europeans) |
| **Calibration** | NI |

| | |
|---|---|
| **Study** | Song et al., 2022 |
| **Transition definition and method of diagnosis** | NI |
| **Time of outcome occurrence or summary of duration of follow-up:** | 2 years |
| **Method for selection of predictors for inclusion in multivariable modelling** | Univariate screening. |
| **Method for selection of predictors during multivariable modelling** | L1 regularisation. |
| **Shrinkage of predictor weights or regression coefficients** | L1 regularisation |
| **Discrimination (95% CI)** | C-index 0.98 (NI) |
| **Calibration** | NI |

| | |
|---|---|
| **Study** | Tavares et al., 2022 |
| **Transition definition and method of diagnosis** | Transition to psychosis during the follow-up period was established according to the Diagnostic and Statistical Manual of Mental Disorders, Fourth Edition (DSM-IV) criteria based on clinical consensus between at least two experienced psychiatrists. |
| **Time of outcome occurrence or summary of duration of follow-up:** | 2+ years |
| **Method for selection of predictors for inclusion in multivariable modelling** | Pre-specification of dimension reduction. |
| **Method for selection of predictors during multivariable modelling** | N/A |
| **Shrinkage of predictor weights or regression coefficients** | Elastic Net |
| **Discrimination (95% CI)** | Balanced Accuracy 46.2% (40.5, 52.4) |
| **Calibration** | NI |

| | |
|---|---|
| **Study** | Zhang et al., 2022 |
| **Transition definition and method of diagnosis** | Conversion to psychosis, determined using the criteria for Presence of Psychotic Symptoms from SIPS CHR-converters were defined by the presence of a 6-level positive symptom (the rating "6" refers to severe and psychotic) that was either dangerous, disorganized, or occurring at least an hour a day on average over four days a week, for at least 16 hours total. |
| **Time of outcome occurrence or summary of duration of follow-up:** | 1 year |
| **Method for selection of predictors for inclusion in multivariable modelling** | Pre-specified. |
| **Method for selection of predictors during multivariable modelling** | N/A |
| **Shrinkage of predictor weights or regression coefficients** | N/A |
| **Discrimination (95% CI)** | C-index 0.68 (0.53-0.82) |
| **Calibration** | NI |

| | |
|---|---|
| **Study** | Zhang et al., 2023a |
| **Transition definition and method of diagnosis** | Conversion to psychosis, determined using the criteria for Presence of Psychotic Symptoms from SIPS CHR-converters were defined by the presence of a 6-level positive symptom (the rating "6" refers to severe and psychotic) that was either dangerous, disorganized, or occurring at least an hour a day on average over four days a week, for at least 16 hours total. |
| **Time of outcome occurrence or summary of duration of follow-up:** | 1 year |
| **Method for selection of predictors for inclusion in multivariable modelling** | Pre-specified. |
| **Method for selection of predictors during multivariable modelling** | N/A |
| **Shrinkage of predictor weights or regression coefficients** | N/A |
| **Discrimination (95% CI)** | C-index 0.897 (0.75 - 1.04) |
| **Calibration** | NI |

| | |
|---|---|
| **Study** | Zhang et al., 2023b |
| **Transition definition and method of diagnosis** | The presence of a six-level positive symptom on the SIPS that was either dangerous, disorganized, or occurred at least 1h a day on average over four days a week for at least 16h in total. |
| **Time of outcome occurrence or summary of duration of follow-up:** | 1 years |
| **Method for selection of predictors for inclusion in multivariable modelling** | Univariate AUROC. |
| **Method for selection of predictors during multivariable modelling** | N/A |
| **Shrinkage of predictor weights or regression coefficients** | N/A |
| **Discrimination (95% CI)** | C-index 0.73 (0.58-0.87) |
| **Calibration** | NI |

### Supplementary Table 3: PROBAST Results

| **Publication reference** | Bousman et al., 2018 | Chan et al., 2015 | Clark et al., 2016 | Dickens et al., 2021 | Jeffries et al., 2016 | Koutsouleris et al., 2021 | Lavoie et al., 2017 | Li et al., 2022 | Mongan et al., 2020 | Perkins et al., 2020 | Perkins et al., 2017 | Song et al., 2022 |
|---|---|---|---|---|---|---|---|---|---|---|---|---|
| **Models of interest** | rs12621220 model | Validation in prodrome | Fatty acid model | Prediction of transition | Leukocyte miRNA model | PRS-based model | Erythrocyte gluthatione model | Prediction of transition | Original model | Model with PRS score | Blood analyte model | Conversion model |
| 1.1 Were appropriate data sources used, e.g. cohort, RCT or nested case-control study data? | Y | Y | Y | Y | Y | Y | Y | Y | Y | Y | Y | Y |
| 1.2 Were all inclusions and exclusions of participants appropriate? | NI | NI | N | Y | PN | PY | N | NI | Y | PN | PN | NI |
| **Risk of bias introduced by selection of participants** | **Unclear** | **Unclear** | **Unclear** | **Low** | **Unclear** | **Low** | **Unclear** | **Unclear** | **Low** | **Unclear** | **Unclear** | **Unclear** |
| **Concern that the included participants and setting do not match the review question** | **Unclear** | **Unclear** | **Unclear** | **Low** | **Unclear** | **Low** | **Unclear** | **Unclear** | **Low** | **Unclear** | **Unclear** | **Unclear** |
| 2.1 Were predictors defined and assessed in a similar way for all participants? | PY | Y | Y | Y | PY | Y | Y | Y | Y | PN | PY | Y |
| 2.2 Were predictor assessments made without knowledge of outcome data? | NI | Y | NI | NI | NI | NI | Y | NI | Y | NI | Y | NI |
| 2.3 Are all predictors available at the time the model is intended to be used? | Y | Y | Y | Y | Y | Y | Y | Y | Y | Y | Y | Y |
| **Risk of bias introduced by predictors or their assessment** | **Unclear** | **Low** | **Unclear** | **Unclear** | **Unclear** | **Unclear** | **Low** | **Unclear** | **Low** | **Unclear** | **Low** | **Unclear** |
| **Concern that the definition, assessment or timing of predictors in the model do not match the review question** | **Low** | **Low** | **Low** | **Low** | **Unclear** | **Low** | **Low** | **Unclear** | **Unclear** | **Low** | **Low** | **Unclear** |
| 3.1 Was the outcome determined appropriately? | PY | Y | PY | PY | PY | Y | PY | PY | PY | PY | Y | NI |

| | | | | | | | | | | | | |
|---|---|---|---|---|---|---|---|---|---|---|---|---|
| 3.2 Was a pre-specified or standard outcome definition used? | PY | PY | Y | NI | Y | Y | Y | PY | NI | Y | Y | NI |
| 3.3 Were predictors excluded from the outcome definition? | Y | Y | Y | PY | Y | Y | Y | Y | PY | PY | Y | PY |
| 3.4 Was the outcome defined and determined in a similar way for all participants? | Y | PY | Y | N | Y | Y | Y | Y | N | Y | Y | NI |
| 3.5 Was the outcome determined without knowledge of predictor information? | PY | PY | NI | PY | PY | Y | Y | PY | PY | PY | Y | NI |
| 3.6 Was the time interval between predictor assessment and outcome determination appropriate? | PY | NI | PY | PY | Y | Y | PY | PY | PY | Y | Y | PY |
| **Risk of bias introduced by the outcome or its determination** | **Low** | **Unclear** | **Unclear** | **Unclear** | **Low** | **Low** | **Low** | **Low** | **Unclear** | **Low** | **Low** | **Unclear** |
| **Concern that the outcome, its definition, timing or determination do not match the review question** | **Low** | **Unclear** | **Low** | **Low** | **Low** | **Low** | **Low** | **Low** | **Low** | **Low** | **Low** | **Low** |
| 4.1 Were there a reasonable number of participants with the outcome? | PN | PN | N | N | N | PN | PN | PN | PN | PN | N | N |
| 4.2 Were continuous and categorical predictors handled appropriately? | Y | Y | N | Y | Y | Y | Y | Y | Y | PY | Y | Y |
| 4.3 Were all enrolled participants included in the analysis? | NI | NI | N | NI | N | N | N | NI | N | N | N | N |
| 4.4 Were participants with missing data handled appropriately? | NI | PN | PY | NI | NI | PY | N | N | PY | PY | NI | NI |
| 4.5 Was selection of predictors based on univariable analysis avoided? | N | | N | N | N | Y | Y | N | Y | Y | N | N |
| 4.6 Were complexities in the data (e.g. censoring, competing risks, sampling of controls) accounted for appropriately? | NI | NI | PN | NI | NI | PN | NI | NI | PN | PN | NI | NI |

| | | | | | | | | | | | | |
|---|---|---|---|---|---|---|---|---|---|---|---|---|
| 4.7 Were relevant model performance measures evaluated appropriately? | N | N | N | N | N | N | N | N | N | N | N | N |
| 4.8 Were model overfitting and optimism in model performance accounted for? | N | | N | PN | PN | Y | N | PN | Y | Y | PN | PN |
| 4.9 Do predictors and their assigned weights in the final model correspond to the results from multivariable analysis? | PY | | Y | Y | Y | Y | Y | Y | PY | Y | PY | NI |
| **Risk of bias introduced by the analysis** | **High** | **High** | **High** | **High** | **High** | **High** | **High** | **High** | **High** | **High** | **High** | **High** |
| **Overall judgement of risk of bias** | **High** | **High** | **High** | **High** | **High** | **High** | **High** | **High** | **High** | **High** | **High** | **High** |
| **Overall judgement of applicability** | **Unclear** | **Unclear** | **Unclear** | **Low** | **Unclear** | **Low** | **Unclear** | **Unclear** | **Unclear** | **Unclear** | **Unclear** | **Unclear** |

**cont.**

| **Publication reference** | Mondelli et al., 2023 | Tavares et al., 2023 | Zhang et al., 2022 | Zhang et al., 2023a | Zhang et al., 2023b |
|---|---|---|---|---|---|
| **Models of interest** | Prediction of transition | eQTL model | Conversion model | Conversion model | Conversion model |
| 1.1 Were appropriate data sources used, e.g. cohort, RCT or nested case-control study data? | Y | Y | Y | Y | Y |
| 1.2 Were all inclusions and exclusions of participants appropriate? | Y | PY | PY | PY | NI |
| **Risk of bias introduced by selection of participants** | **Low** | **Low** | **Low** | **Low** | **Unclear** |
| **Concern that the included participants and setting do not match the review question** | **Low** | **Low** | **Low** | **Low** | **Unclear** |
| 2.1 Were predictors defined and assessed in a similar way for all participants? | Y | PN | Y | Y | Y |
| 2.2 Were predictor assessments made without knowledge of outcome data? | Y | NI | NI | NI | NI |
| 2.3 Are all predictors available at the time the model is intended to be used? | Y | Y | Y | Y | Y |
| **Risk of bias introduced by predictors or their assessment** | **Low** | **Unclear** | **Unclear** | **Unclear** | **Unclear** |
| **Concern that the definition, assessment or timing of predictors in the model do not match the review question** | **Low** | **Low** | **Low** | **Low** | **Low** |
| 3.1 Was the outcome determined appropriately? | PY | Y | Y | Y | Y |

| | | | | | |
|---|---|---|---|---|---|
| 3.2 Was a pre-specified or standard outcome definition used? | NI | Y | Y | Y | Y |
| 3.3 Were predictors excluded from the outcome definition? | PY | Y | PY | PY | PY |
| 3.4 Was the outcome defined and determined in a similar way for all participants? | N | Y | Y | Y | Y |
| 3.5 Was the outcome determined without knowledge of predictor information? | PY | PY | PY | PY | PY |
| 3.6 Was the time interval between predictor assessment and outcome determination appropriate? | PY | Y | Y | Y | Y |
| **Risk of bias introduced by the outcome or its determination** | **Unclear** | **Low** | **Low** | **Low** | **Low** |
| **Concern that the outcome, its definition, timing or determination do not match the review question** | **Low** | **Low** | **Low** | **Low** | **Low** |
| 4.1 Were there a reasonable number of participants with the outcome? | PN | PN | N | N | N |
| 4.2 Were continuous and categorical predictors handled appropriately? | Y | PY | Y | Y | Y |
| 4.3 Were all enrolled participants included in the analysis? | N | N | N | N | N |
| 4.4 Were participants with missing data handled appropriately? | PN | PY | PN | PN | NI |
| 4.5 Was selection of predictors based on univariable analysis avoided? | Y | Y | N | N | N |

| | | | | | |
|---|---|---|---|---|---|
| 4.6 Were complexities in the data (e.g. censoring, competing risks, sampling of controls) accounted for appropriately? | NI | PN | NI | NI | NI |
| 4.7 Were relevant model performance measures evaluated appropriately? | N | N | N | N | N |
| 4.8 Were model overfitting and optimism in model performance accounted for? | Y | Y | N | N | N |
| 4.9 Do predictors and their assigned weights in the final model correspond to the results from multivariable analysis? | PY | Y | NI | NI | NI |
| **Risk of bias introduced by the analysis** | **High** | **High** | **High** | **High** | **High** |
| **Overall judgement of risk of bias** | **High** | **High** | **High** | **High** | **High** |
| **Overall judgement of applicability** | **Low** | **Low** | **Low** | **Low** | **Unclear** |
